# The impact of chronic pain on brain gene expression

**DOI:** 10.1101/2024.05.20.24307630

**Authors:** Lily Collier, Carina Seah, Emily M. Hicks, Traumatic Stress Brain Research Group, Paul E. Holtzheimer, John H. Krystal, Matthew J. Girgenti, Laura M. Huckins, Keira J.A. Johnston

## Abstract

**Background:** Chronic pain affects one fifth of American adults, contributing significant public health burden. Chronic pain mechanisms can be further understood through investigating brain gene expression.

**Methods:** We tested differentially expressed genes (DEGs) in chronic pain, migraine, lifetime fentanyl and oxymorphone use, and with chronic pain genetic risk in four brain regions (dACC, DLPFC, MeA, BLA) and imputed cell type expression data from 304 postmortem donors. We compared findings across traits and with independent transcriptomics resources, and performed gene-set enrichment.

**Results:** We identified two chronic pain DEGs: B4GALT and VEGFB in bulk dACC. We found over 2000 (primarily BLA microglia) chronic pain cell type DEGs. Findings were enriched for mouse microglia pain genes, and for hypoxia and immune response. Cross-trait DEG overlap was minimal.

**Conclusions:** Chronic pain-associated gene expression is heterogeneous across cell type, largely distinct from that in pain-related traits, and shows BLA microglia are a key cell type.

## Introduction

Chronic pain affects roughly one fifth of adults in the United States ^1,2^, and chronic pain conditions (including low back pain and headache disorders) rank in the global top 10 non-infectious diseases contributing to disability adjusted life years ^3^. Chronic pain is defined as pain lasting at least three months ^4,5^. It is associated with a wide range of physical conditions and it may follow surgery, injury or physical trauma ^6,7^. Mechanisms of chronic pain development are not fully understood, and for many individuals treatment is not effective, particularly long term ^8–10^.

Understanding the etiopathology of chronic pain requires that researchers investigate and compare the roles of predisposition to pain development, and effects stemming from experience of pain. Studies of predisposition including large genome-wide association studies have uncovered hundreds of trait-associated variants and genes ^11,12^, and have suggested various Central Nervous System (CNS) pathways to be important in chronic pain ^13^. However, these findings (as with GWAS findings more broadly) have yet to be translated into full mechanistic understanding of disease development, or into actionable treatment. One step toward addressing this knowledge gap is through analysis at the gene expression and transcriptomic level, intermediate steps between genotype (GWAS findings) and phenotype (chronic pain). We and others have applied transcriptomic imputation approaches to translate GWAS findings into gene-tissue associations ^14,15^, identifying brain regions with a putative role in chronic pain including hippocampus, cerebellum, amygdala and frontal cortex among others. However, we do not yet know which brain regions and cell types are primarily involved with chronic pain development. Rodents studies have identified >700 differentially expressed genes in brain and spinal cord, associated with a range of phenotypes including chronic pain, post-surgical pain, neuropathic pain, and spinal injury ^16–20^.

Meanwhile, human gene expression studies have focused on identifying genes associated with specific chronic pain conditions, including fibromyalgia ^21^, osteoarthritis ^22^, migraine ^23^, sickle cell disease ^24^, endometriosis ^25^, and others ^26–31^. However, sample sizes tend to be small, and typically do not include brain tissue. Here, we present the largest study of differentially expressed genes of brain tissue in chronic pain to date. We assess gene expression in post-mortem brain tissues from 304 human donors, from four brain regions previously implicated in pain processing and chronic pain (basal and medial amygdala, prefrontal and anterior cingulate cortex), with detailed phenotype data. We ask whether chronic pain has region- and cell-type specific transcriptomic signatures in the human brain. Defining chronic pain is complex, and it is vital to differentiate pain-associated transcriptomic signal from genes associated with predisposition, specific pain-causing conditions (such as migraine), or genes with expression changes resulting from use of pain-alleviating medications (such as opioids). Using donor phenotype and genotype data, we determine whether the transcriptomic signatures identified in our analyses arise from predisposition to chronic pain, experience of pain itself, or due to treatments or medications taken to alleviate pain (e.g., fentanyl, oxymorphone).

## Methods

### Postmortem brain dataset description

We obtained data for 304 postmortem human brains, donated at time of autopsy through US medical examiner’s offices, from the Veteran’s Association (VA) National PTSD Brain Bank and Traumatic Stress Brain Research group (**Table 1**). Retrospective clinical diagnostic review of toxicology and next-of-kin interviews were performed in this previous study, and a wide range of clinical, anthropometric, psychiatric and life history information, including on trauma history, drug use, and presence of chronic pain, was collected. 115 total donors were female, 189 were male, with a mean age of death of 46.75 years.

Bulk tissue was sampled from four regions of each brain; the dorsolateral anterior cingulate cortex (dACC), dorsolateral prefrontal cortex (DLPFC), basolateral amygdala (BLA), and medial amygdala (MeA), totaling 1216 samples. Collection of brain tissue samples postmortem, genotyping, and producing gene expression data for this cohort is described in detail elsewhere^32,33^.

### Chronic pain phenotype

Chronic pain in this dataset was encoded as present/absent (1/0) by researchers associated with the original data collection, and may include a wide range of chronic pain conditions, locations of chronic pain on the body, and varying severity. Chronic pain presence/ absence was derived from multiple data sources including health records and next of kin interview during retrospective clinical diagnostic review. Migraine, fentanyl lifetime use, and oxymorphone lifetime use were determined using the same approach.

### Imputation of cell-type level gene expression data from bulk tissue

To obtain imputed cell type level gene expression, we carried out cell type deconvolution and subsequent estimation of cell type level gene expression. Since choice of reference panel impacts cell type imputation accuracy ^34^, we used two reference panels. We imputed both cortical bulk tissues (DLPFC, dACC) with PsychENCODE reference cell types ^35–37^, and both amygdala bulk tissues (BLA, MeA), using an amygdala specific reference panel ^38^ (see Supplement). Six cell types were available in both reference datasets (endothelial cells, microglia, inhibitory neurons, excitatory neurons, oligodendrocytes and astrocytes) (see also Supplement).

Imputation was carried out in the same way across all tissues. First, using the bulk RNA-seq raw counts, we constructed a matrix of transcripts, using CIBERSORTx ^39^ and the appropriate reference panel (cortex or amygdala) to estimate cell type proportions for bulk tissue regions. bMind ^40^ was used to impute cell type-specific gene expression for a total of six cell types in each region (astrocytes, endothelial cells, excitatory neurons, inhibitory neurons, microglia, and oligodendrocytes).

### Finding differentially expressed genes using bulk tissue and cell type level expression data

We calculated surrogate variables (SVs) using bulk tissue gene expression data and preserving for chronic pain, in order to capture sources of variability not directly measured in the study ^41^ using R package sva ^42^. We then checked for significant correlation between these SVs and our measured variables using Pearson correlation tests. We next removed variables with more than half of samples showing missing data, and then retained variables that were not correlated with at least one SV (representing variables that were not fully captured by SVs). We then checked for collinearity amongst these retained variables, removing variables and re-checking correlations in a stepwise manner until a set of non-correlated (non-collinear) variables remained. These uncorrelated-with-SVs, non-collinear variables were included as covariates alongside our SVs and chronic pain phenotype in a linear regression model to find differentially expressed genes in chronic pain (see Table S1 for regression model per bulk tissue/ cell type analysis). We applied Bonferroni correction within-tissue (total 4 tissues) to our regression p value results. We then repeated DEG analysis steps (SVA, SV correlation check, collinearity check, DEG regression analysis) for cell type level gene expression data, applying Bonferroni correction within tissue within cell type (total 24 sets) to our results.

We repeated DEG analyses for lifetime fentanyl use, lifetime oxymorphone use, and migraine in both bulk tissue and cell type. We carried out correlation tests among all lifetime opioid use variables – fentanyl was significantly (p < 0.05, Pearson correlation test) correlated with most other lifetime opioid use variables, except oxymorphone. Therefore, we chose fentanyl and oxymorphone for DEG analyses, in order to investigate whether any chronic pain DEGs we found reflected opioid treatment or use as a result of chronic pain.

### Multiple Test Corrections

Unless stated otherwise, multiple test correction was carried out in each DEG analysis using a Bonferroni correction for the number of genes tested. For bulk tissue analyses we applied Bonferroni correction within-tissue, and for cell type analyses we applied Bonferroni multiple testing correction within-tissue within cell type. We also apply a secondary experiment-wise multiple test correction (see Supplement).

### Pathway analyses of chronic pain DEGs using FUMA

We carried out gene-set enrichment tests within FUMA ^43^ using significant genes (P_Bonferroni_ < 0.05). We required at least 10 unique significant genes for inclusion in this analysis. Taking DEGs from cell-type analyses in each tissue (total N=24 initial lists of significant genes), we included gene lists containing more than ten unique genes. We used as background 17,550 genes that were included in each of our DEG analyses per cell type, available in FUMA, and assigned an Ensembl gene ID.

### Comparing cell type DEGs and DAM/ARM and McGill Transcriptomic Pain Signatures data

We next compiled a list of differentially expressed genes from the ‘transcriptomics pain signatures database’ (TSPdb) maintained by the human pain genetics lab at McGill University ^44^, where data is available for human whole blood, synovial fluid, and cartilage transcriptomics experiments. We retained genes from human experiments in whole blood (the majority of the human experiments in this database 1,260,387 (76% of experiments)), where the contrast investigated was ‘pain vs no pain’, where sequencing was through high throughput and not microarray, and where experiments had both males and females included. This resulted in a list of 203,984 experimental results comprised of 23,499 unique genes, 15,968 of which are also tested in each of our cell type DEG analyses. We carried out a series of Fisher’s exact tests for enrichment of each list of chronic pain genes per cell type in this McGill database list, with the list of 15,968 shared genes as background. Note that it was not possible to directly test for DEG enrichment (i.e., identical gene-tissue results) as opposed to enrichment for genes implicated in DEGs (regardless of tissue), as the only human tissues available in the McGill database are non-brain.

We then compiled a list of genes from the same database from experiments on mice (Mus musculus) and in brain and nervous tissues (Medical Subject Headings (MeSH) beginning ‘A08..’) (2,940,318 experiments). We then again included results where contrast investigated was pain vs no pain, experiments used high throughput sequencing, and including both males and females (206,827 experiments). The final database subset contained results for four tissues: microglia (A08.637.400), brain stem (A08.186.211.132), spinal ganglia (A08.800.350.340), and sciatic nerve (A08.800.800.720.450.760). We carried out a series of Fisher’s exact tests for enrichment of each list of chronic pain genes per cell type in each set of database results per tissue.

We also compared our chronic pain microglia DEGs to disease-associated microglia (DAM) and activation response microglia (ARM) genes, using gene lists from publicly available information on DAMs and ARMs from two studies ^45,46^ compiled by Thrupp et al ^47^ (Thrupp et al Table S1 ‘Genesets previous studies’) (360 genes, of which 349 were also tested in our analyses). We did not carry out formal enrichment tests as it was not possible to discern the total number of genes shared as background between our analyses and the Thrupp et al gene list experiments.

### Polygenic risk score analyses

We carried out analyses to uncover DEGs associated with polygenic risk of (proxying predisposition to) chronic pain, using GWAS summary statistics for multisite chronic pain (MCP) ^48^.

First, using PLINK 1.9 and 2.0 ^49^ we carried out recommended quality control (QC) procedure for PRSice2 PRS analyses ^50^. We removed duplicate and ambiguous (palindromic) SNPs from our GWAS summary statistics, as well as SNPs with a low minor allele frequency (MAF < 0.01) and low imputation quality (< 0.8).

Next, we carried out QC steps for genotype data associated with the postmortem brain dataset. We excluded SNPs with MAF < 0.01, duplicate SNPs, SNPs with missing genotype call rates > 0.01, and SNPs not in Hardy-Weinberg equilibrium (p < 1×10^-6^). We also removed samples with missing call rates greater than 0.01. Next, we LD-pruned SNPs in order to obtain a set of SNPs in linkage equilibrium, using window size 200 bp, step size 25 bp, and a correlation threshold of 0.25 and PLINK –indep-pairwise. Then, we calculated sample heterozygosity using PLINK 2.0 – het function, and excluded samples with values greater than 3 SDs from the mean sample heterozygosity value. We then used PLINK 1.9 –check-sex function to flag samples where genetic sex did not match reported sex, which were then removed. We filtered for genetic relatedness using PLINK 1.9 –rel-cutoff at a threshold of π > 0.125 (3^rd^ degree relatives). We performed PCA analysis using PLINK pca and the QC’d genotype data to obtain the first 20 genetic principal components for inclusion in the PRS calculation as covariates, to account for population structure (particularly differing genetic ancestry between source GWAS cohort and postmortem brain tissue donors).

We then used PRSice2 to calculate MCP-PRS for these N=250 post-QC postmortem brain cohort participants. PRSice2 runs multiple PRS analyses at varying GWAS p value thresholds for inclusion, returning a best-fit PRS (measured by PRS-trait regression R2 value). All SNPs passing QC and associated with MCP were included in the PRS calculation. DEG analyses were then carried out as previously described using cell type level expression data, with the trait of interest being MCP-PRS value for each donor rather than chronic pain.

## Results

### Chronic pain impacts the brain transcriptome in a region- and cell-type specific manner

In order to identify genes associated with chronic pain in cortical (dACC, DLPFC) and amygdala (medial and basolateral) brain regions, we performed a differential expression analysis comparing bulk tissue from individuals with and without histories of chronic pain. Two genes were significantly downregulated in the dACC in individuals with chronic pain compared to controls (**Figure 1A**), B4GALT2 (p=1.45×10^-6^, β= –0.936), and VEGFB (p=1.2×10^-7^, β =– 0.945).

**Figure 1:**
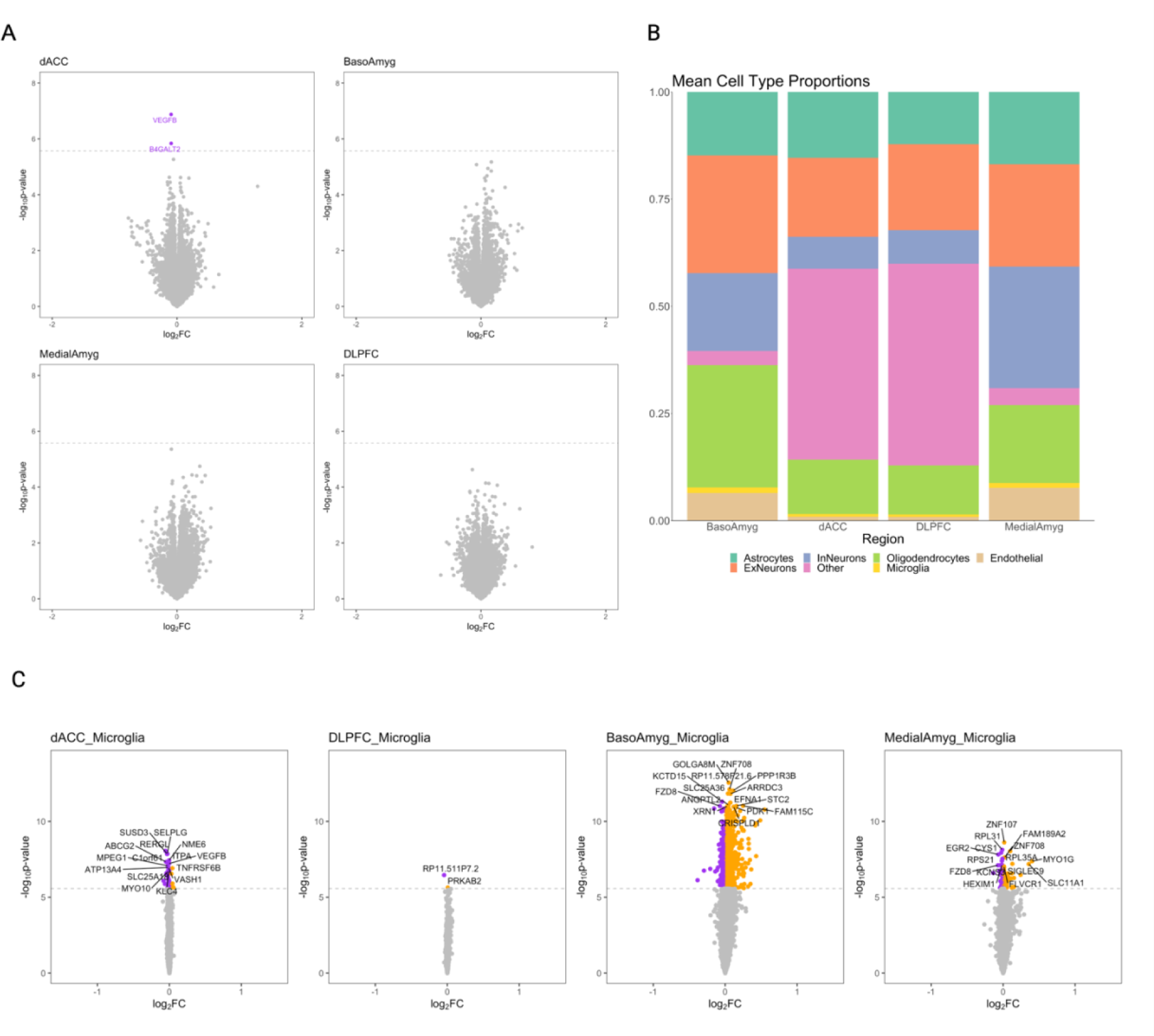
Chronic pain DEGs are found in the dACC and in microglia of the basolateral amygdala in bulk and cell-type-level analyses. 1A: Bulk tissue chronic pain DEGs. Purple: significantly (P_Bonferroni_ < 0.05) downregulated, dotted line = DEG regression p value significance threshold for that bulk region. FC = fold change. 1B: Imputed cell type proportions vary across bulk regions – note neurons only present in reference data for cortex (Psychencode) and oligodendrocyte progenitor cells (OPCs) only present in amygdala reference data (Yu et al), OPCs and Neurons both marked ‘Other’ in this figure panel. FC = fold change. 1C: Chronic pain cell type DEGs in microglia per region. Purple = significantly (P_Bonferroni_ < 0.05) downregulated, orange = significantly (P_Bonferroni_ < 0.05) upregulated, dotted line = p value significance threshold. For legibility only the top 15 DEGs are labeled in cell type results panel. FC = fold change. InN = inhibitory neuron, ExN = excitatory neuron.

Since bulk tissue gene expression likely represents a combination of signal across cell types sampled in each region, we repeated DEG analyses for each brain region using deconvoluted, imputed cell type level expression data, across six cell types (**Figure 1B**; astrocytes, endothelial cells, excitatory neurons, inhibitory neurons, microglia, and oligodendrocytes). 2093 unique genes were significantly associated with chronic pain in these imputed cell types, with the majority of associations (1810/2326) in BLA microglia of the (**Figure 1C, Figure S1**). Given the relatively small proportions of microglia in these brain regions, these associations represent a significant enrichment over what might be expected by chance (p_binomial_ < 1×10^-50^). There was also a significant enrichment of associations in medial amygdala endothelial cells (N DEGs = 254, p_binomial_ = 1.3×10^-42^). Findings in amygdala region oligodendrocyte progenitor cells (OPCs) are presented in the Supplement (**Table S2**).

### Chronic pain associated genes are enriched in hypoxia response, ribosome component, and immunity and infection-related pathways

We explored potential functional impact of chronic pain DEGs through gene set enrichment analyses, including all cell type DEG results with a sufficient number of unique significant genes; dACC microglia, dACC oligodendrocytes, BLA microglia, MeA microglia, MeA astrocytes, MeA endothelial, and MeA oligodendrocytes. Chronic pain associated genes in across these tissues and cell types were enriched for a total of 140 pathway enrichments comprising 101 unique pathways (p_Bonferroni_ = 1.53×10^-6^, adjusting across cell and tissue types, **Table S3, Supplement**).

### Significant enrichment of chronic pain genes in transcriptomic experiment results for mouse CNS but not human whole blood

Chronic pain genes differentially expressed in the MeA microglia were significantly (Fisher’s p = 0.01) enriched for mouse spinal ganglia genes from the McGill TSPdb (**Table 2**). Chronic pain genes DE in the MeA astrocytes, and in dACC microglia and endothelial cells were also significantly enriched for mouse microglia pain genes. There was no significant enrichment of mouse sciatic nerve or mouse brain stem genes in our chronic pain DEG findings.

Sixty-seven genes identified as significantly DE in microglia were also up/down-regulated in DAMs and/or ARMs (**Table 3**). Of these, 13 show concordant direction of effect across chronic pain, DAMs, and ARMs, 21 show concordant effect across chronic pain and DAMs, and 17 across chronic pain and ARMs. None of our chronic pain genes were enriched for pain results from human whole blood (Fisher’s p > 0.05).

### Chronic pain and polygenic risk for Multisite Chronic Pain are associated with differential expression in unique sets of genes

To assess whether chronic pain DEGs represent predisposition to chronic pain, or are associated with downstream impacts of pain on brain gene expression, we sought to identify DEGs associated with polygenic risk for multisite chronic pain (MCP) at the cell type level. We found 21 DEGs significantly associated with MCP-PRS (**Table 4**), none of which were identified as chronic pain DEGs in our previous analyses.

### Migraine and chronic pain DEGs significantly overlap in medial amygdala endothelial cells

In order to distinguish genes associated with chronic pain from genes associated with specific chronic pain conditions, we tested for migraine DEGs in bulk- and imputed cell type data. Although no genes were significantly associated with migraine in bulk tissue, 942 DEGs were associated at the cell-type level, with an over-representation in microglia (754/942 DEGs, p_binomial_ < 2×10^-16^; albeit in the MeA rather than BLA), in dACC oligodendrocytes (52/942, binomial p = 0.027), and in MeA endothelial cells (117/942, binomial p = 3.3×10^-25^). Twelve DEGs were associated with both migraine and chronic pain (Table S4), representing a significant overlap overall (Fisher’s p = 0.004), with this enrichment being driven solely by DEGs in MeA endothelial cells (hypergeometric p = 0.02, Table S5).

### Chronic pain associations are not driven by opioid use

Chronic pain gene associations may also be confounded by long-term use of medications taken to address pain. To test this, we identified DEGs associated with lifetime fentanyl (fentanyl DEGs) and oxymorphone use (oxymorphone DEGs), and compared both single-gene and genome-wide associations to our chronic pain signatures.

At the bulk level, we identified one gene significantly downregulated in individuals with a lifetime history of oxymorphone use in MeA (GPR158.AS1; p=1.17×10^-06^). This DEG was not a chronic pain DEG in bulk tissue. At the cell-type level, we identified 744 significant oxymorphone DEGs, of which 13 were also previously associated with chronic pain (Table S6, Figure 2). Oxymorphone DEGs significantly overlapped with chronic pain DEGs (hypergeometric p < 0.05) in both BLA and MeA endothelial cells and microglia (Table S7). No genes were significantly associated with fentanyl use in bulk tissue. Cell-type specific analyses identified 17 significant fentanyl DEGs (Table S8); however, none of these DEGs overlapped with chronic pain.

**Figure 2:**
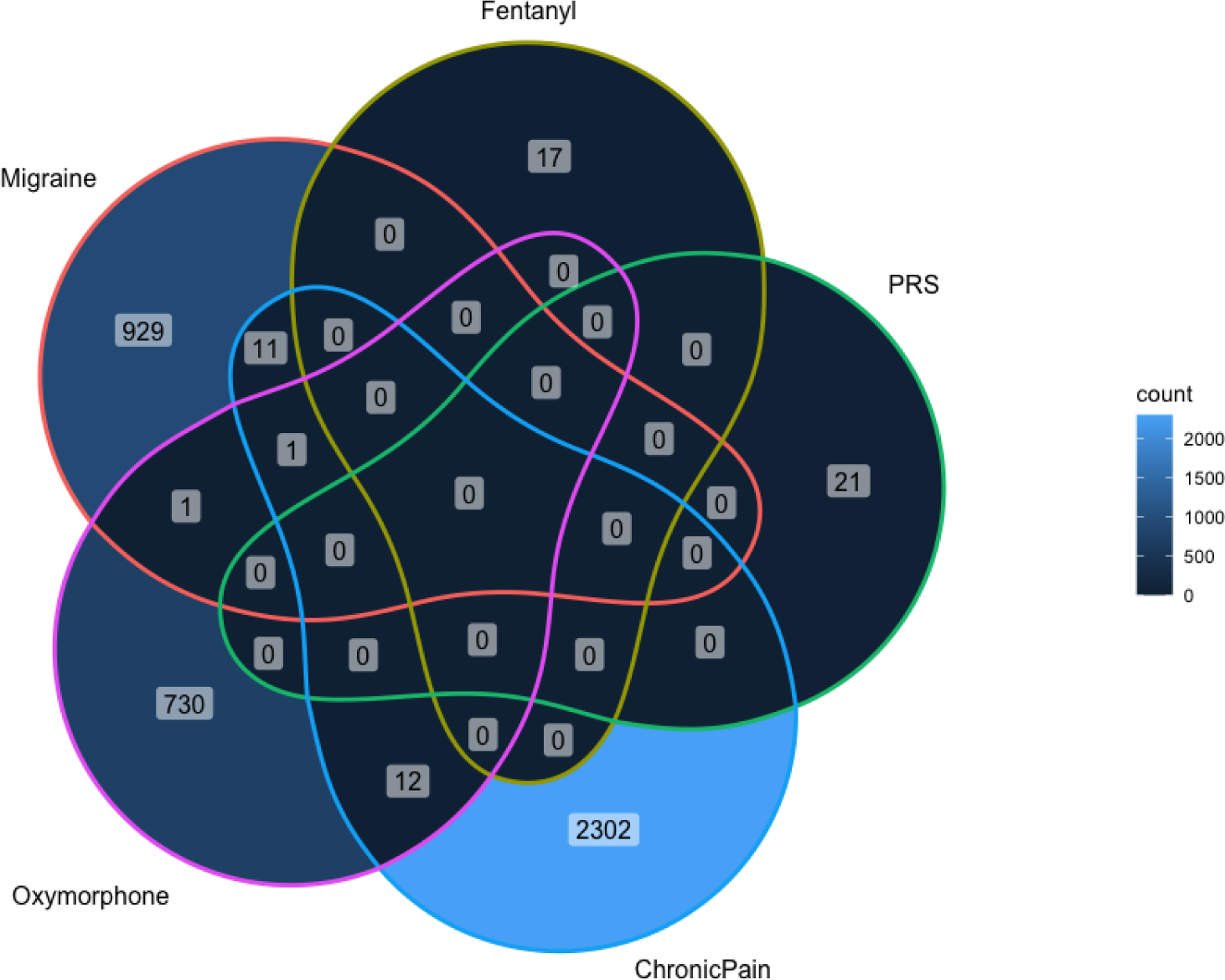
Cell type DEG overlap across all traits (lifetime oxymorphone use, lifetime fentanyl use, multisite chronic pain PRS, chronic pain, and migraine) tends to be low. PRS – polygenic risk score (MCP-PRS).

## Discussion

Understanding the etiopathology of chronic pain requires careful dissection of chronic pain risk, experience, and treatment. Here, we characterize the impact of each of these distinct factors on brain gene expression; we show that chronic pain primarily impacts gene expression in amygdala microglia, and that this transcriptomic signature is for the most part distinct from the signatures of pain predisposition, or the impacts of taking pain-alleviating medications. Our study is the largest to examine the impact of chronic pain across brain regions through differential expression analysis of brain tissue. Strengths of our study include large sample size, with 304 donors and 4 brain regions (all previously implicated in chronic pain) sampled per donor. In addition, a large amount of phenotypic data for each donor is available (160+ variables on lifestyle, trauma, psychiatric and medical diagnoses, and lifetime substance and medication use), allowing us to thoroughly account for sources of variance in gene expression that are not due to chronic pain or other trait-of-interest status. We use two separate region matched reference datasets for cortex and amygdala to study cell type level gene expression imputation. In addition, unusually for chronic pain studies, our sample is almost perfectly balanced in terms of reported sex in donors with chronic pain.

### Microglia represent a key cell type in chronic pain

We leverage advances in cell-type imputation, coupled with large, well-powered cell imputation reference panels to expand our study beyond region specificity, to look at cell-type specificity. We find the majority of our associations in microglia. In addition, we find our chronic pain genes to be enriched in mouse results in CNS and microglia from the pain transcriptomic signatures database, and to show partial overlap with genes associated with DAMs (disease-activated microglia) and ARMs (activated response microglia), reactive microglia phenotypes associated with neurodegenerative disease ^46,51,52^.

In the context of chronic pain, microglia have previously been implicated in imaging studies of low back pain patients ^53^, in the transition from acute to chronic pain in rodent models of spinal nerve transection ^54^, in chronic visceral pain ^55^, and in general neuroplasticity related to chronic pain ^56^. Specifically in BLA microglia, studies in rodents found inhibition of these cells increased anti-nociceptive effects of opioid drugs ^57^, and reduced pain-induced depressive behaviour in a bone cancer pain model ^58^. Our results indicate that BLA microglia play an important part in human chronic pain, that significant overlap is present between human and mouse chronic pain DEGs in microglia, and that the transcriptomic changes in microglia in chronic pain do not exactly match many of those seen in DAMs/ARMs.

### Impact of pain-related factors and conditions on gene expression in the brain

Importantly, our study compares and contrasts different factors involved in pain. There may be key differences in transcriptional changes that occur as a result of living with chronic pain, with predisposition to developing chronic pain, and with indirect results of chronic pain such as potential opioid use. Furthermore, distinct genes may be differentially expressed in chronic pain compared to more specific chronic pain conditions such as migraine.

### Genetic predisposition to chronic pain is transcriptomically distinct from experience of chronic pain

We found distinct sets of genes associated with experience of chronic pain (i.e., observed in brain expression) compared to genetic predisposition to chronic pain (which we measure through polygenic risk for Multisite Chronic Pain). We note that although our sample size (304 donors) is low for polygenic risk score analyses, this sample size paired with the large base GWAS (N > 380, 000 participants), is considered acceptably powered ^59^.

There are no DEGs shared between polygenic risk for multisite chronic pain and chronic pain as a trait. In contrast to chronic pain DEGs, DEGs for polygenic risk of multisite chronic pain were found primarily in dACC oligodendrocytes (19/21 DEGs), in contrast to BLA microglia which appear to play a role in established chronic pain. Previous studies have shown oligodendrocytes may contribute to chronic pain development ^60^, in addition to evidence in rodent models that oligodendrocyte ablation can induce central pain without immune / other glial cell input ^61^.

Genes differentially expressed with increasing polygenic risk of MCP included genes with previous evidence of involvement in pain and cardiac phenotypes, including Fam13b (associated with mouse cardiac phenotypes, 82), TRAF5 (vascular inflammation and atherosclerosis, 83), and FAM184B (heart disease and hip pain ^64^). These associations are in line with established links between chronic pain and heart conditions ^65^, including in our own previous work showing pain gene associations with cardiac dysrhythmias in a large EHR ^14^. Other MCP-PRS associated genes have previous evidence of association with pain phenotypes, including DOCK3 ^66^, LAMB1 ^67^, DUSP4 ^68^, and C1QTNF7 ^69^. Several genes were associated with nerve injury and neuropathies, including NCOA5 ^70^, EPHA5 ^71^, FZD4 ^72^, and MRPL48 ^73^, and NFKBID ^74^.

### Opioid and chronic pain DEG overlap is small and concentrated in the amygdala

At the cell level, none of the chronic pain DEGs overlapped with those associated with lifetime fentanyl use, and 13 DEGs overlapped between chronic pain and lifetime oxymorphone use. These 13 overlapping DEGs represented significant general enrichment of chronic pain DEGs within oxymorphone DEGs (Fisher’s p = 0.0001), but this was driven by four specific cell types (each showing hypergeometric p < 0.05); BLA microglia, BLA endothelial cells, MeA microglia and MeA endothelial cells.

Overlapping DEGs in oxymorphone and chronic pain had concordant direction of effect (i.e., genes significantly downregulated in chronic pain tend to also be downregulated in lifetime oxymorphone use). Although we did not explicitly examine opioid induced hyperalgesia (OIH), these results suggest transcriptomic changes associated with both increased pain and oxymorphone use are present in these cell types of the amygdala, potentially representing relevant genes and cell types in OIH.

Genes differentially expressed in both oxymorphone and chronic pain (13 DEGs) included those implicated in TNF response and previously linked to Lyme disease (YBX3, ^75,76^), encoding cytokines (CCL2, ^77^), Raine syndrome and bone mineralization (FAM20C, ^78,79^), autocrine signaling and lipid storage (HILPDA, ^80^), connective tissue biogenesis (LOXL2, ^81^), neurogenesis (RASF10, ^82^), and Alzheimer disease, Parkinson disease, schizophrenia, and PTSD

(SERPINA3, ^83–86^). YBX3 gene expression in the brain has also been previously linked to PTSD^87^.

### Migraine and chronic pain DEG overlap is small and driven by amygdala endothelial cells

Pathophysiology of migraine is not completely understood ^88,89^, and as with all conditions where chronic pain is a main symptom, there may be distinct mechanisms underlying condition-specific processes and potential tissue damage vs a general chronic pain phenotype. We explored this through comparing genes differentially expressed in migraine and in chronic pain, finding small (12 DEGs) but significant overlap driven by endothelial cells of the medial amygdala.

Endothelial cells interface with vasculature, and endothelial dysfunction has been previously implicated in migraine, although it is not fully established whether this dysfunction is a cause or consequence of migraine ^90^. Significant overlap in chronic pain and migraine DEGs in medial amygdala endothelial cells could therefore mean transcriptomic changes at endothelial cells and migraine-related pain are specifically linked (as opposed to non-pain related migraine symptoms, and non-migraine/ general chronic pain).

In addition, VEGF, encoded by VEGFA, is a ‘protective angiogenic factor’ in migraine ^90^. VEGFB (in contrast to VEGFA) is ‘dispensable’ in new blood vessel growth in normal tissue but seems to play a more essential role in neurodegenerative diseases and stroke (protecting various vasculature during disease progression) ^91^. VEGF has also been implicated in rodent and human studies of neuroplasticity, stress response, and antidepressant action ^92–96^. Our results showed VEGFB was downregulated in bulk dACC tissue in chronic pain, indicating VEGFB may represent a protective angiogenic factor outside of migraine, and in a general chronic pain context.

### Limitations

There are some key limitations of our study that should be borne in mind when interpreting our results. The chronic pain phenotype is extremely broad and dichotomized. Opioid use phenotypes were similarly broad, and lifetime use did not delineate between prescribed/ non-prescribed, and current or past use. When comparing lifetime fentanyl use and lifetime oxymorphone use, ‘case’ numbers are higher for fentanyl, but far fewer DEGs are found – there are also no overlapping DEGs between the two traits. This may reflect different clinical uses of these drugs, with fentanyl as a more discrete rather than chronic exposure, making effects on gene expression more difficult to capture post-mortem and potentially long after a brief exposure.

Another limitation is lack of human brain tissue experiment data available in the McGill TSPdb for direct comparisons with our results, although we explored enrichment in human whole blood and mouse brain and nervous system results.

### Conclusions

Our results suggest that chronic pain impacts gene expression in a heterogeneous way within brain regions depending on cell type, and signal from cell type expression changes may be hidden in bulk tissue analyses due to heterogeneous cell type populations. We highlight brain region and cell type specific genes differentially expressed in chronic pain. In particular, our results suggest BLA microglia are a key cell type in chronic pain, and that pain related transcriptomic changes are distinct from those seen in neurodegenerative disease and general neuroinflammation. Our microglia findings were also enriched for genes differentially expressed in mouse microglia and spinal ganglia samples, but not in human whole blood, again emphasizing region and cell specificity and suggesting a degree of cross-species conservation of certain chronic pain associated transcriptomic changes. Pathways enriched for genes significantly differentially expressed in these cells in chronic pain include those involved in immune processes and hypoxia response. Migraine, opioid use, genetic predisposition to chronic pain, and chronic pain are all largely distinct at the transcriptome level. Points of overlap, though overall consisting of a small number of genes, can inform potential mechanisms underlying shared characteristics between traits. Differences at the transcriptome level between predisposition to chronic pain and chronic pain can be potentially used to inform tailoring of treatment to stage in chronic pain development.

## Data Availability

Full summary data from DEG analyses results is available upon reasonable request to the authors. Requests for access to raw and processed genotype, gene expression, and phenotype data can be made by researchers to the VA National PTSD Brain Bank https://www.research.va.gov/programs/tissue_banking/PTSD/default.cfm

## Acknowledgements

The authors acknowledge support by the National PTSD Brain Bank of the National Center for PTSD (Department of Veterans Affairs). LMH acknowledges funding from NIMH (R01MH124839, R01MH118278, R01MH125938, RM1MH132648, R01MH136149), NIEHS (R01ES033630), and the Department of Defense (TP220451). JHK acknowledges support from the Clinical Neuroscience Division of the National Center for PTSD (Department of Veterans Affairs). CS acknowledges funding from NIH (F30MH132324).

The authors thank members of the Traumatic Stress Brain Research Group (Consortia Authors): Victor E. Alvarez, MD, David Benedek, MD, Alicia Che, PhD, Dianne A. Cruz, MS, David A. Davis, PhD, Matthew J. Girgenti, PhD, Ellen Hoffman, MD, PhD, Paul E. Holtzheimer, MD, Bertrand R. Huber, MD, PhD, Alfred Kaye, MD, PhD, John H. Krystal, MD, Adam T. Labadorf, PhD, Terence M. Keane, PhD, Mark W. Logue, PhD, Ann McKee, MD, Brian Marx, PhD, Mark W. Miller, PhD, Crystal Noller, PhD, Janitza Montalvo-Ortiz, PhD, Meghan Pierce, PhD,William K. Scott, PhD, Paula Schnurr, PhD, Krista DiSano, PhD, Thor Stein, MD, PhD, Robert Ursano, MD, Douglas E. Williamson, PhD, Erika J. Wolf, PhD, Keith A. Young, PhD.

## Notes

### Competing Interest Statement

J.H.K. has consulting agreements (less than US$10,000 per year) with the following: Aptinyx, Inc. Biogen, Idec, MA, Bionomics, Limited (Australia), Boehringer Ingelheim International, Epiodyne, Inc., EpiVario, Inc., Janssen Research & Development, Jazz Pharmaceuticals, Inc., Otsuka America Pharmaceutical, Inc., Spring Care, Inc., Sunovion Pharmaceuticals, Inc.; is the co-founder for Freedom Biosciences, Inc.; serves on the scientific advisory boards of Biohaven Pharmaceuticals, BioXcel Therapeutics, Inc. (Clinical Advisory Board), Cerevel Therapeutics, LLC, Delix Therapeutics, Inc., Eisai, Inc., EpiVario, Inc., Jazz Pharmaceuticals, Inc., Neumora Therapeutics, Inc., Neurocrine Biosciences, Inc., Novartis Pharmaceuticals Corporation, PsychoGenics, Inc., Takeda Pharmaceuticals, Tempero Bio, Inc., Terran Biosciences, Inc..; has stock options with Biohaven Pharmaceuticals Medical Sciences, Cartego Therapeutics, Damona Pharmaceuticals, Delix Therapeutics, EpiVario, Inc., Neumora Therapeutics, Inc., Rest Therapeutics, Tempero Bio, Inc., Terran Biosciences, Inc., Tetricus, Inc.; and is editor of Biological Psychiatry with income greater than $10,000

### Author Declarations

Ethical approval associated with VA Biorepository Brain Bank: The National PTSD Brain Bank/ IRB 32887/ IRBNet ID 1578200. Consent was provided by next of kin at time of brain donation. We also note the research in our study is not human subjects research under HHS 45 CFR part 46, a human subject is defined as a living individual about whom an investigator conducting research obtains data through intervention or interaction with the individual or identifiable private information.This research involved the analysis of gene expression (RNA) data, along with associated phenotype, and genotype data, from postmortem human brain tissue.

