## Supplement for "The impact of chronic pain on brain gene expression"

**Cell-type deconvolution with an alternate reference dataset**

We repeated cell type deconvolution of expression data from our bulk tissue samples with CIBERSORTx [1], using an alternate reference dataset downloaded from Gene Expression Omnibus[2] (Series GSE195445) and associated with a set of RNA-seq experiments using amygdala samples only [3]. First the Yu et al human dataset Seurat[4] object was processed to produce the files required for CIBERSORTx web application signature matrix generation step (columns of samples and rows of genes, with expression values expressed in TPM). Conversion to TPM using DGEobj.utils function 'convertCounts' requires gene lengths, which were obtained using biomaRt[5] and querying using gene symbols ('external_gene_name') to find start and end positions of each gene. This produced a reference expression dataset for 15, 387 genes from three human donors, each with seven cell types tested (astrocytes, endothelial cells, excitatory neurons, inhibitory neurons, microglia, oligodendrocytes, and oligodendrocyte progenitor cells). Then we performed CIBERSORTx step 2, using the generated signature matrix and Girgenti et al postmortem bulk tissue expression data in TPM. We then calculated the mean cell type proportion for each brain region, and compared this with cell type proportion means using our previous reference data set as described in the main manuscript.

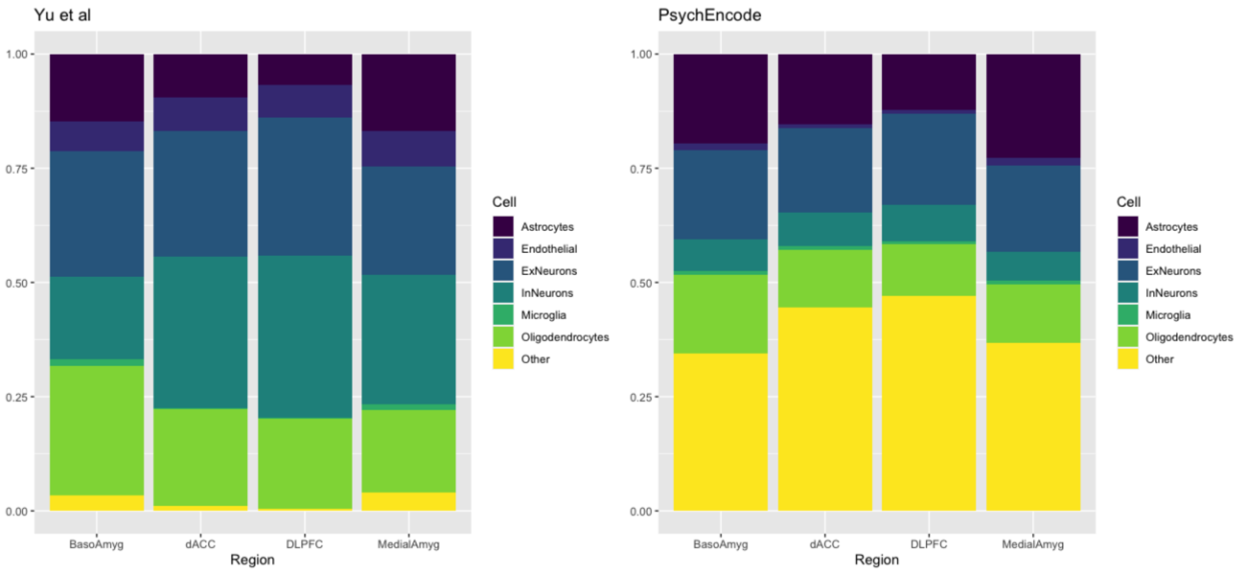


Cell type proportions across the four brain regions following cell-type expression imputation using amygdala (Yu et al) and cortex (psychENCODE) reference panels. Note: 'Other' in Yu et al is OPC, in PsychEncode 'Neurons'.

Significant differences in cell type proportions are seen, particularly in overall neuron content. We opted to impute cell type level expression data in cortical and amygdala bulk data separately using the two distinct reference data sets (psychENCODE cortical samples for dACC and DLPFC, and Yu et al amygdala samples for MeA and BLA).

**Additional DEG analysis multiple-testing correction**

We consider the five separate sets (PRS, oxymorphone, fentanyl, chronic pain, and migraine) of DEG analyses not to require additional cross-experiment multiple testing correction, i.e., multiple testing correction as described in the main manuscript is sufficient. However, we applied an additional, even more stringent DEG regression p value threshold (0.05 / number of genes tested within that tissue and cell type / 5 (number of traits in total across entire study)). While this does reduce the number of significant DEGs identified, a t-test comparing N DEGs at the original p value threshold vs. additional correction threshold p = 0.49, and in particular the main bulk of our chronic pain findings remain in the BLA microglia (original p value threshold N DEG in BLA microglia: 1810 (77% of all chronic pain DEGs), with additional correction: 1260 (82%)).

**FUMA gene-set enrichment p value threshold**

In order to calculate a Bonferroni-adjusted p value threshold we downloaded the entirety of the gene sets used in the latest version of FUMA GENE2FUNC from the FUMA website 'GENE2FUNC_genesets_v156plus.tar.gz'. We then removed duplicated gene sets and sets that were nested (gene sets for FUMA are sourced from MSigDB [6] and all are included despite e.g., certain sets being repeated due to their classification as both a canonical pathway and a reactome pathway (child category of canonical pathway)). This results in a total of 32, 584 independent gene sets and therefore a Bonferroni p value threshold of 0.05/32,584 within each tissue-cell type gene-set enrichment (pathway) analysis.

**DEGs in Oligodendrocyte Progenitor Cells (OPCs)**

We find 18 significant ($P_{Bonferroni} < 0.05$) chronic pain DEGs, 47 fentanyl DEGs, 1 oxymorphone DEG, 3 migraine DEGs, and 0 PRS-DEGs in MeA OPCs. We find 4 significant chronic pain DEGs, 64 fentanyl DEGs, 3 oxymorphone DEGs, 0 migraine DEGs, and 0 PRS-DEGs in BLA OPCs.

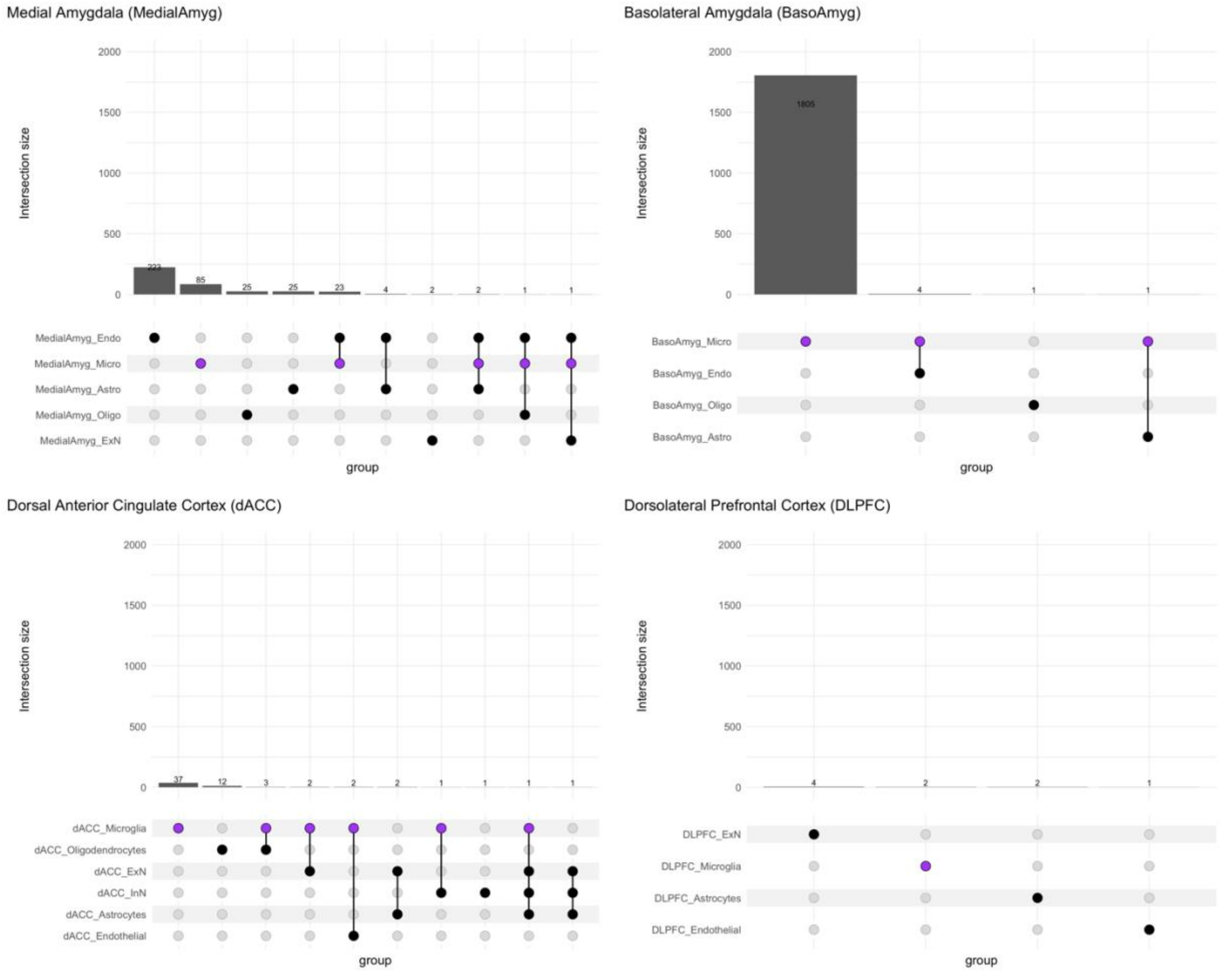


**Fig S1: Cell type DEGs in chronic pain – comparison across regions and cell types.** Majority of findings are in BLA microglia, and number of DEGs shared between cell types is low.

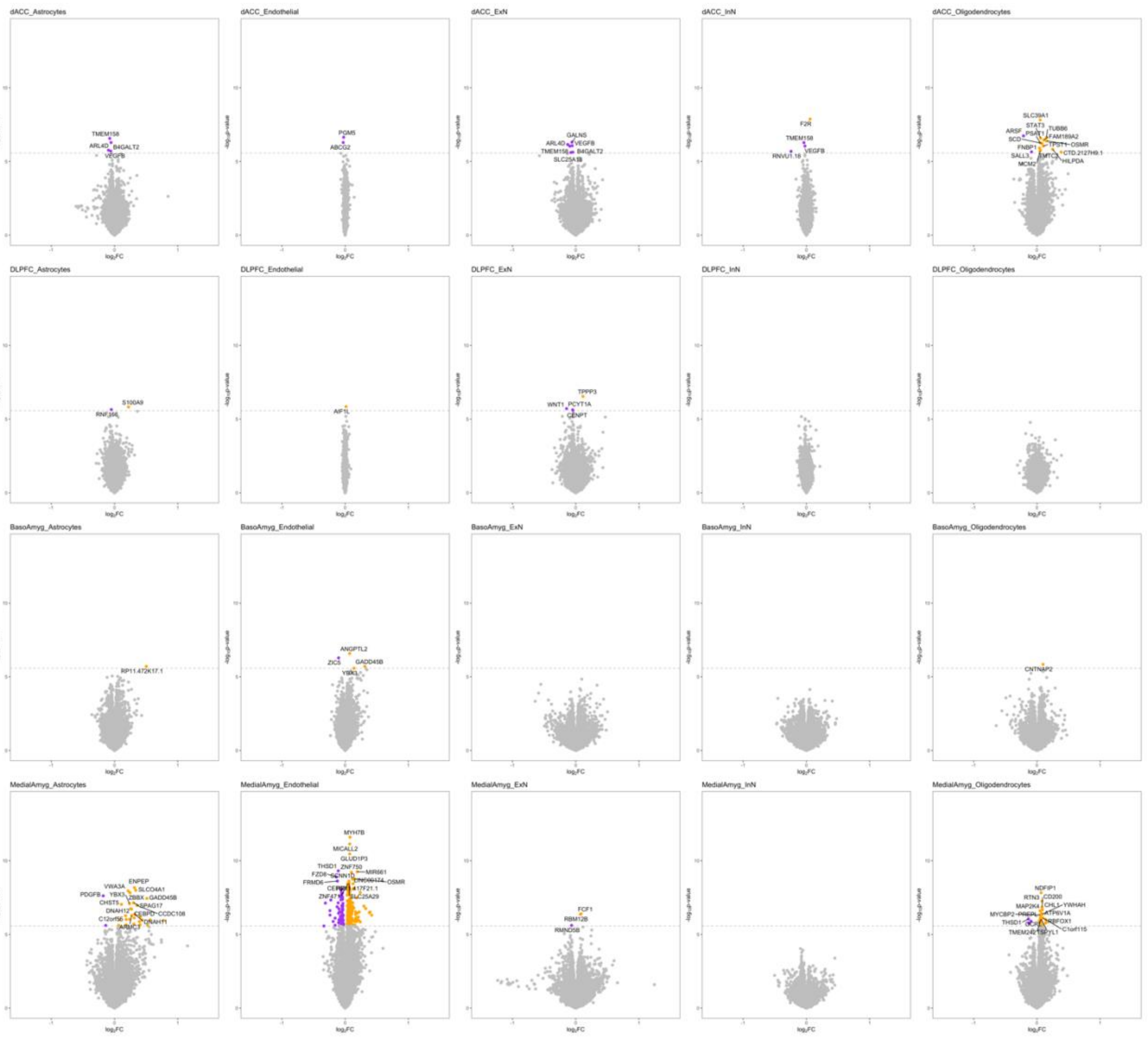


**Fig S2: Additional genes differentially expressed across cell types in chronic pain.** Chronic pain cell type DEGs in cell types per region. Purple = significantly ($P_{Bonferroni} < 0.05$) downregulated, orange = significantly ($P_{Bonferroni} < 0.05$) upregulated, dotted line = p value significance threshold. For legibility only the top 15 DEGs are labeled. FC = fold change. InN = inhibitory neuron, ExN = excitatory neuron.

**Table S1: DEG analyses per-gene regression models**. BLA = Basolateral Amygdala, MeA = Medial Amygdala, InN = inhibitory neuron, ExN = excitatory neuron, dACC = dorsal anterior cingulate cortex, DLPFC = dorsolateral prefrontal cortex, SV = surrogate variable, model = DEG regression model fitted per gene. In model column; 'gene' = gene expression value, 'trait' = trait value (case/control, yes/no for oxymorphone lifetime use, fentanyl lifetime use, migraine, chronic pain, quantitative MCP-PRS value for PRS).

| Trait | Analysis type | Region | Cell | N SVs | model |
|---|---|---|---|---|---|
| Chronic Pain | Bulk | DLPFC | NA | 22 | gene ~ trait + SVs + Handedness + Migraine + Amphetamines |
| | | dACC | NA | 21 | gene ~ trait + SVs + Handedness + Tramadol + Amphetamines + Ketamine + Anticholinergics..Antiparkinsonians + personal_count |
| | | BLA | NA | 25 | gene ~ trait + SVs + Manner_of_death_Suicide + Delta.9.THC.Active + Barbiturates + witness_any |
| | | MeA | NA | 27 | gene ~ trait + SVs + PTSD_PrimaryDx + Barbiturates + personal_count |
| | Cell-type | BLA | InN | 53 | gene ~ trait + SVs + Acetone + Cocaethylene + Benzoylecgonine_mg_L_blood + Hydrocodone + Ketamine + Anticholinergics..Antiparkinsonians + Hallucinogens + ECT |
| | | | ExN | 43 | gene ~ trait + SVs + Isopropanol + Famhx.Suicide + combat_only_count |
| | | | Oligodendrocytes | 44 | gene ~ trait + SVs + X11.Hydroxy.Delta.9.THC.Active + Amphetamines + Barbiturates + Ketamine + personal_any |
| | | | Endothelial | 38 | gene ~ trait + SVs + Race + BMI..calculated. + X11.Hydroxy.Delta.9.THC.Active + Barbiturates + disaster_count + assault_count |
| | | | Astrocytes | 49 | gene ~ trait + SVs + Isopropanol + X6.AM + Tramadol + |

| | | | | | |
|---|---|---|---|---|---|
| | | | | | Anticholinergics..Antiparkinsonians + Hallucinogens + EA_UA_neglect_count + personal_count |
| | | | Microglia | 28 | gene ~ trait + SVs + Handedness + BMI..calculated. + Isopropanol + X11.Hydroxy.Delta.9.THC.Active + Benzoylecgonine + Barbiturates + Ketamine + Anticholinergics..Antiparkinsonians + Hallucinogens + SA_count + witness_any |
| | | dACC | InN | 39 | gene ~ trait + SVs |
| | | | ExN | 40 | gene ~ trait + SVs + Cocaethylene + Amphetamines + childhood_count |
| | | | Oligodendrocytes | 55 | gene ~ trait + SVs + Cocaethylene + Benzoylecgonine_mg_L_blood + Nicotine_ng_mL + Barbiturates + Anticholinergics..Antiparkinsonians |
| | | | Endothelial | 21 | gene ~ trait + SVs + Cocaethylene + Nicotine_ng_mL + Amphetamines + Barbiturates + disaster_count + childhood_count |
| | | | Astrocytes | 43 | gene ~ trait + SVs + Cocaethylene + Amphetamines + assault_count |
| | | | Microglia | 12 | gene ~ trait + SVs + Handedness + Acetone + Ethanol_mg_dL + X11.Hydroxy.Delta.9.THC.Active + Benzoylecgonine + Hydrocodone + Barbiturates + Anticholinergics..Antiparkinsonians + Past.Self.Mutilation |
| | | DLPFC | InN | 38 | gene ~ trait + SVs + BMI..calculated. + Cocaethylene + Hallucinogens + witness_count + personal_count |
| | | | ExN | 37 | gene ~ trait + SVs + Cocaethylene + Barbiturates + Hallucinogens + witness_count |
| | | | Oligodendrocytes | 54 | gene ~ trait + SVs + Handedness + |

| | | | | | |
|---|---|---|---|---|---|
| | | | | | Barbiturates + Ketamine + Anticholinergics..Antiparkinsonians + ECT + witness_count + personal_count + combat_occ_count |
| | | | Endothelial | 17 | gene ~ trait + SVs + Manner_of_death_Accident + GERD + X11.Hydroxy.Delta.9.THC.Active + Benzoylecgonine_mg_L_blood + Barbiturates + Delusions + combat_occ_count |
| | | | Astrocytes | 39 | gene ~ trait + SVs + X11.Hydroxy.Delta.9.THC.Active + Amphetamines + personal_count + combat_occ_count |
| | | | Microglia | 8 | gene ~ trait + SVs + Handedness + Manner_of_death_Accident + Isopropanol + X11.Hydroxy.Delta.9.THC.Active + Benzoylecgonine_mg_L_blood + Anticholinergics..Antiparkinsonians + Anti.Inflammatories + Delusions + combat_occ_count |
| | | MeA | InN | 43 | gene ~ trait + SVs + Acetone + X11.Hydroxy.Delta.9.THC.Active + Barbiturates + Ketamine + Hallucinogens + disaster_count + childhood_count |
| | | | ExN | 50 | gene ~trait + SVs + Manner_of_death_Suicide + Isopropanol + Cocaethylene + Ketamine + Anticholinergics..Antiparkinsonians + ECT |
| | | | Oligodendrocytes | 46 | gene ~ trait + SVs + GERD + Acetone + X11.Hydroxy.Delta.9.THC.Active + Benzoylecgonine_mg_L_blood + Barbiturates + Ketamine + Anticholinergics..Antiparkinsonians + witness_count + combat_occ_count |
| | | | Endothelial | 42 | gene ~ trait + SVs + Nicotine_ng_mL + |

| | | | | | |
|---|---|---|---|---|---|
| | | | | | Barbiturates + Hallucinogens + disaster_count |
| | | | Astrocytes | 51 | gene ~ trait + SVs + Cocaethylene + Hallucinogens + Other.Drugs + ECT + witness_count |
| | | | Microglia | 24 | gene ~ trait + SVs + Acetone + Cocaethylene + Benzoylecgonine_mg_L_blood + Barbiturates + Ketamine + Anticholinergics..Antiparkinsonians + Hallucinogens + witness_count |
| Fentanyl | Bulk | DLPFC | NA | 22 | gene ~ trait + SVs + Handedness + Amphetamines + Barbiturates + personal_count + combat_occ_count |
| | | dACC | NA | 21 | gene ~ trait + SVs + Handedness + Delta.9.THC.Active + Tramadol + Amphetamines + Ketamine + personal_count |
| | | BLA | NA | 25 | gene ~ trait + SVs + Delta.9.THC.Active + Barbiturates + Anticholinergics..Antiparkinsonians + witness_any |
| | | MeA | NA | 27 | gene ~ trait + SVs + PTSD_PrimaryDx + Hydrocodone + Barbiturates + personal_count |
| | Cell-type | BLA | InN | 54 | gene ~ trait + SVs + Acetone + X11.Hydroxy.Delta.9.THC.Active + Cocaethylene + Hydrocodone + Ketamine + Anticholinergics..Antiparkinsonians + Antipsychotics + Hallucinogens + ECT + disaster_count |
| | | | ExN | 43 | Gene ~ trait + SVs + Isopropanol + cocaethylene + witness_count + combat_occ_any |
| | | | Oligodendrocytes | 43 | gene ~ trait + SVs + X11.Hydroxy.Delta.9.THC.Active + Amphetamines + Barbiturates + Ketamine + disaster_count |

| | | | | | |
|---|---|---|---|---|---|
| | | | Endothelial | 38 | gene ~ trait + Svs + Handedness + X11.Hydroxy.Delta.9.THC.Active + Barbiturates + Hallucinogens + witness_count + personal_count |
| | | | Astrocytes | 49 | gene ~ trait + SVs + Ketamine + Anticholinergics..Antiparkinsonians + Hallucinogens + ECT + witness_count + personal_count |
| | | | Microglia | 27 | gene ~ trait + SVs + Isopropanol + X11.Hydroxy.Delta.9.THC.Active + Cocaethylene + Barbiturates + Ketamine + Anticholinergics..Antiparkinsonians + Hallucinogens + ECT + witness_count + personal_count |
| | | dACC | InN | 38 | gene ~ trait + SVs + Isopropanol + Anti.Inflammatories + personal_count |
| | | | ExN | 40 | gene ~ trait + SVs + Cocaethylene + Tramadol + personal_count |
| | | | Oligodendrocytes | 55 | gene ~ trait + SVs + Cocaethylene + Benzoylecgonine_mg_L_blood + Hydrocodone + Barbiturates + Anticholinergics..Antiparkinsonians + witness_count |
| | | | Endothelial | 21 | gene ~ trait + SVs + Cocaethylene + Nicotine_ng_mL + Amphetamines + Barbiturates + disaster_any + childhood_count |
| | | | Astrocytes | 43 | gene ~ trait + SVs + Cocaethylene + Amphetamines + Past.Self.Mutilation + Famhx.Suicide + combat_occ_any |
| | | | Microglia | 13 | gene ~ trait + SVs + Handedness + Acetone + Ethanol_mg_dL + X11.Hydroxy.Delta.9.THC.Active + Morphine + Anticholinergics..Antiparkinsonians + witness_count |

| | | | | | |
|---|---|---|---|---|---|
| | | DLPFC | InN | 38 | gene ~ trait + SVs + Cocaethylene + Hallucinogens + witness_count + personal_count |
| | | | ExN | 36 | gene ~ trait + SVs + Cocaethylene + Benzoylecgonine_mg_L_blood + Barbiturates + Hallucinogens + witness_count |
| | | | Oligodendrocytes | 53 | gene ~ trait + SVs + Barbiturates + Ketamine + Anticholinergics..Antiparkinsonians + ECT + witness_count + personal_count + combat_occ_count |
| | | | Endothelial | 17 | gene ~ trait + SVs + GERD + X11.Hydroxy.Delta.9.THC.Active + Cocaethylene + Barbiturates + ECT + witness_count + personal_count + combat_occ_count |
| | | | Astrocytes | 38 | gene ~ trait + SVs + Cocaethylene + Amphetamines + childhood_count |
| | | | Microglia | 8 | gene ~ trait + SVs + Manner_of_death_Accident + Ethanol + Isopropanol + X11.Hydroxy.Delta.9.THC.Active + Anti.Inflammatories + EA_UA_neglect_count + personal_count + combat_only_count |
| | | MeA | InN | 44 | gene ~ trait + SVs + Acetone + Barbiturates + Ketamine + Anticholinergics..Antiparkinsonians + Hallucinogens + ECT + disaster_count |
| | | | ExN | 50 | gene ~ trait + SVs + Isopropanol + Barbiturates + Ketamine + Anticholinergics..Antiparkinsonians + Hallucinogens + EA_UA_neglect_count |
| | | | Oligodendrocytes | 45 | gene ~ trait + SVs + Acetone + X11.Hydroxy.Delta.9.THC.Active + Benzoylecgonine + |

| | | | | | |
|---|---|---|---|---|---|
| | | | | | Amphetamines + Barbiturates + Ketamine Anticholinergics..Antiparkinsonians witness_count |
| | | | Endothelial | 41 | gene ~ trait + SVs + Nicotine_ng_mL + Barbiturates + Hallucinogens + disaster_count + childhood_count |
| | | | Astrocytes | 50 | gene ~ trait + SVs + Cocaethylene + Benzoylecgonine_mg_L_blood + Hallucinogens + ECT + witness_count |
| | | | Microglia | 24 | gene ~ trait + SVs + Handedness + Isopropanol + Barbiturates + Ketamine + Anticholinergics..Antiparkinsonians + Hallucinogens + witness_count + personal_count |
| Oxymorphone | Bulk | DLPFC | NA | 22 | gene ~ trait + SVs + Amphetamines + personal_count + combat_occ_count |
| | | dACC | | 21 | gene ~ trait + SVs + Delta.9.THC.Active + Amphetamines + Ketamine + personal_any |
| | | BLA | | 25 | gene ~ trait + SVs + Delta.9.THC.Active + Anticholinergics..Antiparkinsonians + witness_count |
| | | MeA | | 27 | gene ~ trait + SVs + Barbiturates + Hx.Other.Trauma + |
| | Cell-type | BLA | InN | 54 | gene ~ trait + SVs + Acetone + Delta.9.Carboxy.THC.Inactive + Ketamine + Anticholinergics..Antiparkinsonians + Hallucinogens + ECT + disaster_count |
| | | | ExN | 42 | gene ~ trait + SVs + Isopropanol + Cocaethylene + Famhx.Suicide + combat_only_count |
| | | | Oligodendrocytes | 44 | gene ~ trait + SVs + Amphetamines + Barbiturates + Ketamine + EA_UA_neglect_count + disaster_count |

| | | | | | |
|---|---|---|---|---|---|
| | | | Endothelial | 38 | gene ~ trait + SVs + Handedness + X11.Hydroxy.Delta.9.THC.Active + Barbiturates + Hallucinogens + witness_count + personal_count |
| | | | Astrocytes | 49 | gene ~ trait + SVs + Isopropanol + Ketamine + Anticholinergics..Antiparkinsonians + Hallucinogens + ECT + witness_count + personal_count |
| | | | Microglia | 28 | gene ~ trait + SVs + Handedness + Isopropanol + X11.Hydroxy.Delta.9.THC.Active + Barbiturates + Ketamine + Anticholinergics..Antiparkinsonians + Hallucinogens + ECT + witness_count + personal_count |
| | | dACC | InN | 39 | gene ~ trait + SVs + Isopropanol + Benzoylecgonine_mg_L_blood + Anti.Inflammatories |
| | | | ExN | 40 | gene ~ trait + SVs + Cocaethylene + Amphetamines + personal_count |
| | | | Oligodendrocytes | 55 | gene ~ trait + SVs + Cocaethylene + Benzoylecgonine_mg_L_blood + Barbiturates + Anticholinergics..Antiparkinsonians + witness_count |
| | | | Endothelial | 20 | gene ~ trait + SVs + Isopropanol + Cocaethylene + Nicotine_ng_mL + Amphetamines + Barbiturates + disaster_any + childhood_count |
| | | | Astrocytes | 43 | gene ~ trait + SVs + Cocaethylene + Benzoylecgonine_mg_L_blood + witness_count |
| | | | Microglia | 12 | gene ~ trait + SVs + Handedness + Acetone + Ethanol_mg_dL + X11.Hydroxy.Delta.9.THC.Active + Benzoylecgonine + Hydrocodone + Barbiturates + |

| | | | | | |
|---|---|---|---|---|---|
| | | | | | Anticholinergics..Antiparkinsonians |
| | | DLPFC | InN | 38 | gene ~ trait + SVs + BMI..calculated. + Cocaethylene + Hallucinogens + witness_count + personal_count |
| | | | ExN | 37 | gene ~ trait + SVs + Delta.9.THC.Active + Benzoylecgonine_mg_L_blood + Hallucinogens + witness_count |
| | | | Oligodendrocytes | 54 | gene ~ trait + SVs + Handedness + Barbiturates + Anticholinergics..Antiparkinsonians + witness_count + personal_count + combat_occ_count |
| | | | Endothelial | 17 | gene ~ trait + SVs + Manner_of_death_Accident + Isopropanol + X11.Hydroxy.Delta.9.THC.Active + Amphetamines + Barbiturates + Delusions + personal_count + combat_occ_count |
| | | | Astrocytes | 38 | gene ~ trait + SVs + Amphetamines + personal_count |
| | | | Microglia | 8 | gene ~ trait + SVs + Handedness + Manner_of_death_Accident + Isopropanol + X11.Hydroxy.Delta.9.THC.Active + Anti.Inflammatories + Delusions + personal_count + combat_occ_count |
| | | MeA | InN | 43 | gene ~ trait + SVs + Acetone + Barbiturates + Ketamine + Hallucinogens + disaster_count + childhood_count |
| | | | ExN | 49 | gene ~ trait + SVs + Isopropanol + Barbiturates + Ketamine + Anticholinergics..Antiparkinsonians + EA_UA_neglect_count |
| | | | Oligodendrocytes | 46 | gene ~ trait + SVs + Other.Opiates + Barbiturates + Ketamine + Anticholinergics..Antiparkinsonians + |

| | | | | | |
|---|---|---|---|---|---|
| | | | | | witness_count + personal_count |
| | | | Endothelial | 41 | gene ~ trait + SVs + Handedness + X11.Hydroxy.Delta.9.THC.Active + Barbiturates + Anti.Inflammatories + witness_count + personal_count + combat_occ_count |
| | | | Astrocytes | 51 | gene ~ trait + SVs + Cocaethylene + Benzoylecgonine_mg_L_blood + Hallucinogens + ECT + witness_count |
| | | | Microglia | 24 | gene ~ trait + SVs + Acetone + Cocaethylene + Nicotine_ng_mL + Barbiturates + Ketamine + Anticholinergics..Antiparkinsonians + Hallucinogens + non_combat_count |
| Migraine | Bulk | DLPFC | NA | 22 | gene ~ trait + SVs + Handedness + Amphetamines + personal_count + combat_occ_count |
| | | dACC | | 21 | gene ~ trait + SVs + Handedness + Delta.9.THC.Active + Tramadol + Amphetamines + Ketamine + personal_count |
| | | BLA | | 25 | gene ~ trait + SVs + Delta.9.THC.Active + Barbiturates + Anticholinergics..Antiparkinsonians + witness_count |
| | | MeA | | 27 | gene ~ trait + SVs + PTSD_PrimaryDx + Barbiturates + personal_count |
| | Cell-type | BLA | InN | 54 | gene ~ trait + SVs + Acetone + Cocaethylene + Hydrocodone + Ketamine + Anticholinergics..Antiparkinsonians + Antipsychotics + Hallucinogens + ECT + disaster_count |
| | | | ExN | 43 | gene ~ trait + SVs + Isopropanol + Cocaethylene + Past.Self.Mutilation + combat_occ_any |
| | | | Oligodendrocytes | 43 | gene ~ trait + SVs + X11.Hydroxy.Delta.9.THC.Active + |

| | | | | | |
|---|---|---|---|---|---|
| | | | | | Amphetamines + Barbiturates + Ketamine + disaster_count |
| | | | Endothelial | 38 | gene ~ trait + SVs + Handedness + Acetone + X11.Hydroxy.Delta.9.THC.Active + Barbiturates + Hallucinogens + childhood_count |
| | | | Astrocytes | 49 | gene ~ trait + SVs + Ketamine + Anticholinergics..Antiparkinsonians + Hallucinogens + ECT + witness_count + personal_count |
| | | | Microglia | 28 | gene ~ trait + SVs + Isopropanol + Cocaethylene + Barbiturates + Ketamine + Anticholinergics..Antiparkinsonians + Hallucinogens + ECT + witness_count + personal_count |
| | | dACC | InN | 38 | gene ~ trait + SVs + BMI..calculated. + Isopropanol + Anti.Inflammatories + personal_count |
| | | | ExN | 40 | gene ~ trait + SVs + Cocaethylene + Benzoylecgonine_mg_L_blood + Anti.Inflammatories |
| | | | Oligodendrocytes | 55 | gene ~ trait + SVs + Cocaethylene + Benzoylecgonine_mg_L_blood + Barbiturates + Anticholinergics..Antiparkinsonians |
| | | | Endothelial | 21 | gene ~ trait + SVs + Cocaethylene + Nicotine_ng_mL + Amphetamines + Barbiturates + disaster_count + childhood_count |
| | | | Astrocytes | 43 | gene ~ trait + SVs + Cocaethylene + Benzoylecgonine_mg_L_blood + witness_count + combat_occ_count |
| | | | Microglia | 12 | gene ~ trait + SVs + Isopropanol + X11.Hydroxy.Delta.9.THC.Active + Cocaethylene + Benzoylecgonine_mg_L_blood + Hydrocodone + |

| | | | | | |
|---|---|---|---|---|---|
| | | | | | Barbiturates + Ketamine + Anticholinergics..Antiparkinsonians + witness_count |
| | | DLPFC | InN | 37 | gene ~ trait + SVs + Cocaethylene + Hallucinogens + witness_count + personal_count |
| | | | ExN | 36 | gene ~ trait + SVs + Manner_of_death_Accident + Delta.9.THC.Active + Benzoylecgonine_mg_L_blood + Hallucinogens |
| | | | Oligodendrocytes | 54 | gene ~ trait + SVs + Handedness + Barbiturates + Ketamine + Anticholinergics..Antiparkinsonians + ECT + witness_count + personal_count + combat_occ_count |
| | | | Endothelial | 17 | gene ~ trait + SVs + BMI..calculated. + X11.Hydroxy.Delta.9.THC.Active + Cocaethylene + Tramadol + Barbiturates + personal_count |
| | | | Astrocytes | 38 | gene ~ trait + SVs |
| | | | Microglia | 8 | gene ~ trait + SVs + Handedness + Manner_of_death_Accident + Isopropanol + X11.Hydroxy.Delta.9.THC.Active + Benzoylecgonine_mg_L_blood + Anticholinergics..Antiparkinsonians + Anti.Inflammatories + Delusions combat_only_count |
| | | MeA | InN | 44 | gene ~ trait + SVs + Acetone + Barbiturates + Ketamine + Hallucinogens + disaster_count + childhood_count |
| | | | ExN | 50 | gene ~ trait + SVs + Isopropanol + Ketamine + Anticholinergics..Antiparkinsonians + EA_UA_neglect_count |
| | | | Oligodendrocytes | 46 | gene ~ trait + SVs + GERD + Acetone + X11.Hydroxy.Delta.9.THC.Active + |

| | | | | | |
|---|---|---|---|---|---|
| | | | | | Benzoylecgonine_mg_L_blood + Barbiturates + Ketamine Anticholinergics..Antiparkinsonians witness_count |
| | | | Endothelial | 41 | gene ~ trait + SVs + Barbiturates + Hallucinogens + witness_count + personal_count |
| | | | Astrocytes | 51 | gene ~ trait + SVs + Cocaethylene + Benzoylecgonine_mg_L_blood + Hallucinogens + ECT + witness_count |
| | | | Microglia | 24 | gene ~ trait + SVs + Acetone + Cocaethylene + Nicotine_ng_mL + Barbiturates + Ketamine + Anticholinergics..Antiparkinsonians + Hallucinogens + non_combat_count |
| PRS | Cell-type | BLA | InN | 49 | gene ~ trait + SVs + Acetone + Cocaethylene + Hydrocodone + Ketamine + Anticholinergics..Antiparkinsonians + Antipsychotics + Last.PPD + ECT + disaster_count |
| | | | ExN | 39 | gene ~ trait + SVs + Race + Barbiturates + witness_count + combat_only_count |
| | | | Oligodendrocytes | 37 | gene ~ trait + SVs + Acetone + Amphetamines + Barbiturates + Ketamine + disaster_count + assault_count |
| | | | Endothelial | 34 | gene ~ trait + SVs + Acetone + X11.Hydroxy.Delta.9.THC.Active + Barbiturates + Hallucinogens + disaster_count + childhood_count |
| | | | Astrocytes | 41 | gene ~ trait + SVs + Acetone + Benzoylecgonine_mg_L_blood + Ketamine + Anticholinergics..Antiparkinsonians + Hallucinogens + ECT + witness_count |

| | | | | | |
|---|---|---|---|---|---|
| | | | Microglia | 26 | gene ~ trait + SVs + Acetone + Cocaethylene + Barbiturates + Ketamine + Anticholinergics..Antiparkinsonians + Hallucinogens + Age.Onset.Smoke + ECT + witness_count |
| | | dACC | InN | 33 | gene ~ trait + SVs + Race+ Anti.Inflammatories + personal_count + age_num_first_trauma_infer |
| | | | ExN | 34 | gene ~ trait + SVs + Race + Acetone + Cocaethylene + Amphetamines + Age.Onset.Smoke + Past.Self.Mutilation |
| | | | Oligodendrocytes | 45 | gene ~ trait + SVs + Manner_of_death_Accident + Delta.9.THC.Active + Benzoylecgonine_mg_L_blood + Barbiturates + Anticholinergics..Antiparkinsonians |
| | | | Endothelial | 19 | gene ~ trait + SVs + Race + Acetone + Cocaethylene + Nicotine_ng_mL + Amphetamines + Barbiturates + witness_count + disaster_count |
| | | | Astrocytes | 37 | gene ~ trait + SVs + Race + Acetone + Cocaethylene + Amphetamines + Age.Onset.Smoke + witness_count |
| | | | Microglia | 11 | gene ~ trait + SVs + Handedness + Acetone + Ethanol_mg_dL + Benzoylecgonine_mg_L_blood + Acetaminophen + Ketamine + Anticholinergics..Antiparkinsonians + Hx.Military.Service + witness_any |
| | | DLPFC | InN | 34 | gene ~ trait + SVs + Handedness + Benzoylecgonine_mg_L_blood + Age.Onset.Smoke + witness_any + combat_occ_any |
| | | | ExN | 34 | gene ~ trait + SVs + Race + Cocaethylene + |

| | | | | | |
|---|---|---|---|---|---|
| | | | | | Amphetamines + Barbiturates + personal_count + age_num_first_trauma_infer |
| | | | Oligodendrocytes | 44 | gene ~ trait + SVs + Race + Delta.9.THC.Active + Barbiturates + Ketamine + Anticholinergics..Antiparkinsonians + ECT + witness_count + personal_count |
| | | | Endothelial | 16 | gene ~ trait + SVs + X11.Hydroxy.Delta.9.THC.Active + Cocaethylene + Nicotine_ng_mL + Amphetamines + Barbiturates + Famhx.Suicide + disaster_count |
| | | | Astrocytes | 34 | gene ~ trait + SVs + Acetone + X11.Hydroxy.Delta.9.THC.Active + Cocaethylene + Amphetamines + combat_occ_count + childhood_count |
| | | | Microglia | 8 | gene ~ trait + SVs + Handedness + Ethanol + Acetone + Delta.9.Carboxy.THC.Inactive + Benzoylecgonine_mg_L_blood + Age.Onset.Smoke + Past.Self.Mutilation + ECT + Hx.Combat.Seen |
| | | MeA | InN | 42 | gene ~ trait + SVs + Acetone + Barbiturates + Ketamine + Anticholinergics..Antiparkinsonians + Hallucinogens + ECT + disaster_count |
| | | | ExN | 44 | gene ~ trait + SVs + Ketamine + Anticholinergics..Antiparkinsonians + ECT + combat_occ_count |
| | | | Oligodendrocytes | 41 | gene ~ trait + SVs + X11.Hydroxy.Delta.9.THC.Active + Amphetamines + Barbiturates + Ketamine + Anticholinergics..Antiparkinsonians witness_count personal_count combat_occ_count |

| | | | | | |
|---|---|---|---|---|---|
| | | | Endothelial | 36 | gene ~ trait + SVs + Acetone + X11.Hydroxy.Delta.9.THC.Active + Barbiturates + Hallucinogens + disaster_count + childhood_count |
| | | | Astrocytes | 42 | gene ~ trait + SVs + plate + Cocaethylene + Amphetamines + childhood_count |
| | | | Microglia | 22 | gene ~ trait + SVs + Cocaethylene +Oxymorphone + Barbiturates + Ketamine + Anticholinergics..Antiparkinsonians + Hallucinogens + witness_count |

### Table S2: Additional DEGs in MeA and BLA OPCs.

| Trait | Region | gene | Z | P | P_bonf |
|---|---|---|---|---|---|
| Chronic pain | BLA | YBX3 | -5.1 | 7.69E-07 | 1.44E-02 |
| | | CD84 | -5.81 | 2.29E-08 | 4.27E-04 |
| | | GADD45A | -5.68 | 4.48E-08 | 8.38E-04 |
| | | BAG3 | -5.37 | 2.06E-07 | 3.86E-03 |
| | MeA | DYNLL1 | -4.9 | 1.88E-06 | 3.51E-02 |
| | | PDGFB | -5.51 | 9.82E-08 | 1.83E-03 |
| | | WDR83OS | -5.35 | 2.21E-07 | 4.14E-03 |
| | | ETV3 | -5.23 | 3.87E-07 | 7.23E-03 |
| | | DUSP4 | -5.09 | 7.54E-07 | 1.41E-02 |
| | | EGR2 | -5.59 | 6.73E-08 | 1.26E-03 |
| | | COX16 | -5.05 | 9.32E-07 | 1.74E-02 |
| | | TEC | 4.86 | 2.29E-06 | 4.27E-02 |
| | | PROSC | -5.75 | 2.99E-08 | 5.60E-04 |
| | | UCN | 5.26 | 3.36E-07 | 6.27E-03 |
| | | CRK | -4.94 | 1.52E-06 | 2.84E-02 |
| | | TRIB1 | -5.09 | 7.56E-07 | 1.41E-02 |
| | | ZFP1 | -5.32 | 2.53E-07 | 4.72E-03 |
| | | ZNF511 | -5.04 | 9.87E-07 | 1.84E-02 |
| | | PRC1 | 4.95 | 1.46E-06 | 2.73E-02 |
| | | LBH | -4.96 | 1.45E-06 | 2.70E-02 |
| | | C1orf233 | 5.05 | 9.10E-07 | 1.70E-02 |
| | | GLUD1P3 | 4.95 | 1.51E-06 | 2.83E-02 |
| Oxymorphone | BLA | MASP1 | -4.87 | 2.24E-06 | 4.18E-02 |

| | | | | | |
|---|---|---|---|---|---|
| | | SPATA13 | -4.83 | 2.64E-06 | 4.93E-02 |
| | | RP11.309I15.1 | -4.83 | 2.64E-06 | 4.93E-02 |
| | MeA | LBH | -5.01 | 1.19E-06 | 2.23E-02 |
| Fentanyl | BLA | EPHA3 | 5.26 | 3.68E-07 | 6.88E-03 |
| | | LZTS1 | 5.45 | 1.51E-07 | 2.81E-03 |
| | | SREBF1 | 5.63 | 5.95E-08 | 1.11E-03 |
| | | CACNG5 | 5.23 | 4.37E-07 | 8.16E-03 |
| | | DBC1 | 5.07 | 8.94E-07 | 1.67E-02 |
| | | ITCH | 4.9 | 1.98E-06 | 3.70E-02 |
| | | PGM1 | 5.74 | 3.41E-08 | 6.38E-04 |
| | | SMOX | 4.95 | 1.58E-06 | 2.96E-02 |
| | | CDC23 | 4.98 | 1.38E-06 | 2.58E-02 |
| | | EIF5 | -5.76 | 3.16E-08 | 5.90E-04 |
| | | KIAA0391 | 4.92 | 1.82E-06 | 3.41E-02 |
| | | TMEM87A | 4.98 | 1.37E-06 | 2.56E-02 |
| | | CRYBA1 | 5.73 | 3.75E-08 | 7.00E-04 |
| | | TNFAIP1 | 5.65 | 5.44E-08 | 1.02E-03 |
| | | ARSB | 5.46 | 1.42E-07 | 2.66E-03 |
| | | HES1 | 5.16 | 5.82E-07 | 1.09E-02 |
| | | PRDM2 | -4.95 | 1.57E-06 | 2.94E-02 |
| | | APH1A | 5.5 | 1.17E-07 | 2.18E-03 |
| | | FMO6P | 5.1 | 8.00E-07 | 1.50E-02 |
| | | AKAP1 | 5.32 | 2.76E-07 | 5.15E-03 |
| | | SDC4 | 4.86 | 2.41E-06 | 4.51E-02 |
| | | ABCC10 | -5.05 | 9.95E-07 | 1.86E-02 |
| | | SNAP25 | -5.2 | 4.85E-07 | 9.07E-03 |
| | | MED10 | -4.93 | 1.72E-06 | 3.21E-02 |
| | | RFK | -5.25 | 3.81E-07 | 7.11E-03 |
| | | ITM2C | 4.91 | 1.89E-06 | 3.54E-02 |
| | | MRPS9 | 4.9 | 1.96E-06 | 3.65E-02 |
| | | CCL21 | 5.03 | 1.09E-06 | 2.03E-02 |
| | | GRHPR | 4.91 | 1.89E-06 | 3.54E-02 |
| | | LRRC32 | 5.04 | 1.05E-06 | 1.97E-02 |
| | | TMBIM6 | 5.03 | 1.10E-06 | 2.05E-02 |
| | | C18orf8 | -4.86 | 2.36E-06 | 4.40E-02 |
| | | GTDC2 | 5 | 1.26E-06 | 2.35E-02 |
| | | CDC123 | 4.9 | 1.98E-06 | 3.70E-02 |
| | | PFKM | 5.65 | 5.37E-08 | 1.00E-03 |
| | | WNT7A | 5.03 | 1.08E-06 | 2.01E-02 |

| | | | | | |
|---|---|---|---|---|---|
| | | SSR2 | 5.56 | 8.64E-08 | 1.61E-03 |
| | | CYBB | -5.01 | 1.22E-06 | 2.28E-02 |
| | | ILK | 5.19 | 5.21E-07 | 9.74E-03 |
| | | PBX3 | 5.29 | 3.28E-07 | 6.14E-03 |
| | | ECI1 | 4.93 | 1.74E-06 | 3.26E-02 |
| | | SF1 | -4.87 | 2.23E-06 | 4.17E-02 |
| | | CX3CR1 | -4.94 | 1.65E-06 | 3.08E-02 |
| | | GPR34 | -5.8 | 2.60E-08 | 4.86E-04 |
| | | LRRC20 | 5.1 | 7.97E-07 | 1.49E-02 |
| | | CES3 | -5.02 | 1.14E-06 | 2.14E-02 |
| | | LINC00982 | 4.98 | 1.38E-06 | 2.59E-02 |
| | | CCDC89 | -4.88 | 2.19E-06 | 4.09E-02 |
| | | C1orf194 | -4.98 | 1.37E-06 | 2.56E-02 |
| | | P2RY13 | -5.09 | 8.18E-07 | 1.53E-02 |
| | | SCN5A | 6.53 | 5.25E-10 | 9.81E-06 |
| | | ZNF75D | 4.85 | 2.50E-06 | 4.67E-02 |
| | | STMN3 | 5.25 | 3.94E-07 | 7.37E-03 |
| | | MPZL1 | 5.64 | 5.85E-08 | 1.09E-03 |
| | | DUSP27 | -4.96 | 1.52E-06 | 2.84E-02 |
| | | SNORD115.2 | 5.17 | 5.76E-07 | 1.08E-02 |
| | | FAM71F2 | -5.41 | 1.76E-07 | 3.29E-03 |
| | | RNVU1.18 | -6.15 | 4.18E-09 | 7.82E-05 |
| | | MLLT11 | -5.24 | 4.09E-07 | 7.64E-03 |
| | | AC009227.2 | 5.21 | 4.78E-07 | 8.93E-03 |
| | | RP11.180I22.2 | 5.2 | 5.00E-07 | 9.34E-03 |
| | | RP3.437I16.1 | -5.57 | 8.22E-08 | 1.54E-03 |
| | | USP17L4 | 5.6 | 7.20E-08 | 1.35E-03 |
| | | MYLK.AS2 | 5.18 | 5.38E-07 | 1.00E-02 |
| | MeA | NDUFAF7 | -5.12 | 6.61E-07 | 1.23E-02 |
| | | NLE1 | 4.83 | 2.53E-06 | 4.74E-02 |
| | | POMGNT1 | 5.42 | 1.48E-07 | 2.77E-03 |
| | | RPLP0 | 4.95 | 1.43E-06 | 2.66E-02 |
| | | HIF1A | 5.06 | 8.43E-07 | 1.57E-02 |
| | | PRPF6 | 5.14 | 5.99E-07 | 1.12E-02 |
| | | IDH3B | 5.07 | 8.28E-07 | 1.55E-02 |
| | | NFAT5 | -4.82 | 2.61E-06 | 4.87E-02 |
| | | MYEF2 | -4.92 | 1.65E-06 | 3.09E-02 |
| | | MTCH2 | 5.12 | 6.51E-07 | 1.22E-02 |
| | | LDHB | 6 | 7.51E-09 | 1.40E-04 |

| | | | | | |
|---|---|---|---|---|---|
| | | WASF1 | -4.87 | 2.12E-06 | 3.95E-02 |
| | | SELK | 4.95 | 1.42E-06 | 2.65E-02 |
| | | NKTR | -5.29 | 2.86E-07 | 5.35E-03 |
| | | PRDX1 | 5.82 | 2.01E-08 | 3.76E-04 |
| | | ALDH6A1 | 5.13 | 6.32E-07 | 1.18E-02 |
| | | PKN1 | 4.85 | 2.31E-06 | 4.31E-02 |
| | | MRPS7 | 5.24 | 3.63E-07 | 6.78E-03 |
| | | CASD1 | -5.44 | 1.39E-07 | 2.60E-03 |
| | | EIF3G | 5.53 | 8.71E-08 | 1.63E-03 |
| | | MGAT1 | 4.9 | 1.81E-06 | 3.39E-02 |
| | | RAMP1 | 5.39 | 1.74E-07 | 3.26E-03 |
| | | PEMT | 5.48 | 1.13E-07 | 2.11E-03 |
| | | CDK4 | 5.06 | 8.48E-07 | 1.59E-02 |
| | | ITM2C | 6.48 | 5.44E-10 | 1.02E-05 |
| | | SCRN1 | 5.28 | 3.08E-07 | 5.75E-03 |
| | | CREBZF | -5.31 | 2.61E-07 | 4.87E-03 |
| | | TRIM29 | -5.19 | 4.62E-07 | 8.63E-03 |
| | | ETNK1 | -4.93 | 1.58E-06 | 2.96E-02 |
| | | GALNT1 | 5.03 | 9.94E-07 | 1.86E-02 |
| | | MICU3 | -5.25 | 3.50E-07 | 6.54E-03 |
| | | UBN2 | -5.27 | 3.22E-07 | 6.02E-03 |
| | | FAM69B | 5.5 | 9.92E-08 | 1.85E-03 |
| | | NDUFV1 | 5.33 | 2.30E-07 | 4.30E-03 |
| | | NSMCE1 | 5.25 | 3.46E-07 | 6.47E-03 |
| | | SNUPN | 4.97 | 1.30E-06 | 2.42E-02 |
| | | PHC3 | -6.79 | 9.83E-11 | 1.84E-06 |
| | | COA4 | 5.06 | 8.60E-07 | 1.61E-02 |
| | | PLCD1 | 5.65 | 4.75E-08 | 8.88E-04 |
| | | LIN54 | -5.28 | 3.08E-07 | 5.76E-03 |
| | | TRIM33 | -5.18 | 4.82E-07 | 9.01E-03 |
| | | SNORA38B | -5.23 | 3.92E-07 | 7.32E-03 |
| | | SNORA80 | -5.83 | 1.89E-08 | 3.54E-04 |
| | | C9orf135 | -4.85 | 2.32E-06 | 4.34E-02 |
| | | AC005594.3 | -5.09 | 7.59E-07 | 1.42E-02 |
| | | SNORD15B | -4.88 | 1.96E-06 | 3.67E-02 |
| | | ACY1 | 5.27 | 3.22E-07 | 6.01E-03 |
| Migraine | MeA | SPAG4 | 4.99 | 1.26E-06 | 2.36E-02 |
| | | CCDC91 | -4.84 | 2.51E-06 | 4.69E-02 |
| | | ZDHHC12 | 4.83 | 2.62E-06 | 4.89E-02 |

**Table S3: Pathways (gene sets) enriched for additional chronic pain cell type DEGs**. Gene sets significantly ($p < 1.53 \times 10^{-6}$) enriched for cell type DEGs with minimum N gene overlap = 10. GO = Gene Ontology. No gene sets were found to be significantly enriched for dACC microglia, dACC oligodendrocytes, medial amygdala astrocytes, and medial amygdala endothelial cell type DEGs.

| Region | Cell Type | Category | GeneSet | N_genes | N_overlap | p |
|---|---|---|---|---|---|---|
| MeA | Microglia | GO biological processes | GOBP_CYTOPLASMIC_TRANSLATION | 125 | 14 | $5.16 \times 10^{-14}$ |
| | | | GOBP_PEPTIDE_METABOLIC_PROCESS | 744 | 20 | $4.12 \times 10^{-08}$ |
| | | | GOBP_PEPTIDE_BIOSYNTHETIC_PROCESS | 626 | 18 | $7.91 \times 10^{-08}$ |
| | | | GOBP_AMIDE_BIOSYNTHETIC_PROCESS | 752 | 18 | $1.17 \times 10^{-06}$ |
| | | | GOBP_AMIDE_METABOLIC_PROCESS | 1009 | 21 | $1.28 \times 10^{-06}$ |
| | | Chemical and Genetic pertubation | HSIAO_HOUSEKEEPING_GENES | 332 | 14 | $2.46 \times 10^{-08}$ |
| | | GO cellular component | GOCC_CYTOSOLIC_RIBOSOME | 67 | 13 | $2.57 \times 10^{-16}$ |
| | | | GOCC_RIBOSOME | 180 | 16 | $2.86 \times 10^{-14}$ |
| | | | GOCC_RIBOSOMAL_SUBUNIT | 149 | 15 | $3.04 \times 10^{-14}$ |
| | | | GOCC_RIBONUCLEOPROTEIN_COMPLEX | 668 | 19 | $3.89 \times 10^{-08}$ |
| | | GO molecular function | GOMF_STRUCTURAL_CONSTITUENT_OF_RIBOSOME | 132 | 14 | $1.11 \times 10^{-13}$ |
| | | | GOMF_STRUCTURAL_MOLECULE_ACTIVITY | 591 | 18 | $3.31 \times 10^{-08}$ |
| | | Computational gene sets | MORF_TPT1 | 73 | 13 | $8.45 \times 10^{-16}$ |
| | | | MODULE_83 | 266 | 19 | $5.08 \times 10^{-15}$ |
| | | | MORF_ACTG1 | 105 | 13 | $1.16 \times 10^{-13}$ |
| | | | GNF2_EIF3S6 | 93 | 12 | $6.46 \times 10^{-13}$ |
| | | | MORF_NPM1 | 126 | 13 | $1.27 \times 10^{-12}$ |
| | | | MODULE_114 | 285 | 17 | $2.92 \times 10^{-12}$ |
| | | | MODULE_151 | 269 | 16 | $1.41 \times 10^{-11}$ |
| | | | GNF2_FBL | 117 | 11 | $2.06 \times 10^{-10}$ |
| | | | MORF_NME2 | 126 | 11 | $4.60 \times 10^{-10}$ |
| | | | MODULE_32 | 209 | 11 | $9.32 \times 10^{-08}$ |
| | | Cell type signature | TRAVAGLINI_LUNG_CD4_NAIVE_T_CELL | 109 | 15 | $2.59 \times 10^{-16}$ |

| | | | | | | |
|---|---|---|---|---|---|---|
| | | | RUBENSTEIN_SKELETAL_MUSCLE_T_CELLS | 142 | 15 | $1.47 \times 10^{-14}$ |
| | | | BUSSLINGER_GASTRIC_PPP1R1B_POSITIVE_CELLS | 92 | 13 | $1.98 \times 10^{-14}$ |
| | | | TRAVAGLINI_LUNG_BRONCHIAL_VESSEL_1_CELL | 196 | 16 | $1.09 \times 10^{-13}$ |
| | | | HAY_BONE_MARROW_NAIVE_T_CELL | 321 | 19 | $1.53 \times 10^{-13}$ |
| | | | TRAVAGLINI_LUNG_CLUB_CELL | 92 | 12 | $5.65 \times 10^{-13}$ |
| | | | RUBENSTEIN_SKELETAL_MUSCLE_SATELLITE_CELLS | 260 | 17 | $6.65 \times 10^{-13}$ |
| | | | MURARO_PANCREAS_ACINAR_CELL | 622 | 24 | $7.86 \times 10^{-13}$ |
| | | | RUBENSTEIN_SKELETAL_MUSCLE_B_CELLS | 128 | 13 | $1.56 \times 10^{-12}$ |
| | | | BUSSLINGER_DUODENAL_TRANSIT_AMPLIFYING_CELLS | 161 | 14 | $1.76 \times 10^{-12}$ |
| | | | BUSSLINGER_DUODENAL_DIFFERENTIATING_STEM_CELLS | 257 | 16 | $7.08 \times 10^{-12}$ |
| | | | LAKE_ADULT_KIDNEY_C7_PROXIMAL_TUBULE_EPITHELIAL_CELLS_S3 | 106 | 11 | $7.00 \times 10^{-11}$ |
| | | | BUSSLINGER_DUODENAL_IMMUNE_CELLS | 807 | 24 | $1.82 \times 10^{-10}$ |
| | | | LAKE_ADULT_KIDNEY_C10_THIN_ASCENDING_LIMB | 305 | 15 | $9.17 \times 10^{-10}$ |
| | | | LAKE_ADULT_KIDNEY_C12_THICK_ASCENDING_LIMB | 341 | 15 | $4.21 \times 10^{-09}$ |
| | | | BUSSLINGER_DUODENAL_STEM_CELLS | 260 | 13 | $1.08 \times 10^{-08}$ |
| | | | LAKE_ADULT_KIDNEY_C19_COLLECTING_DUCT_INTERCALATED_CELLS_TYPE_A_MEDULLA | 283 | 13 | $2.96 \times 10^{-08}$ |
| | | | RUBENSTEIN_SKELETAL_MUSCLE_PCV_E | 188 | 11 | $3.15 \times 10^{-08}$ |

| | | | | | | |
|---|---|---|---|---|---|---|
| | | | NDOTHELIAL_CELLS | | | |
| | | | TRAVAGLINI_LUNG_CAPILLARY_INTERMEDIATE_2_CELL | 353 | 14 | $5.29x10^{-08}$ |
| | | | TRAVAGLINI_LUNG_MESOTHELIAL_CELL | 542 | 17 | $5.47x10^{-08}$ |
| | | | LAKE_ADULT_KIDNEY_C9_THIN_ASCENDING_LIMB | 231 | 11 | $2.56x10^{-07}$ |
| | | Canonical Pathways | REACTOME_EUKARYOTIC_TRANSLATION_ELONGATION | 64 | 14 | $2.77x10^{-18}$ |
| | | | KEGG_RIBOSOME | 60 | 13 | $5.42x10^{-17}$ |
| | | | WP_CYTOPLASMIC_RIBOSOMAL_PROTEINS | 62 | 13 | $8.62x10^{-17}$ |
| | | | REACTOME_RESPONSE_OF_EIF2AK4_GCN2_TO_AMINO_ACID_DEFICIENCY | 74 | 13 | $1.02x10^{-15}$ |
| | | | REACTOME_CELLULAR_RESPONSE_TO_STARVATION | 122 | 15 | $1.47x10^{-15}$ |
| | | | REACTOME_NONSENSE_MEDIATED_DECAY_NMD | 83 | 13 | $4.92x10^{-15}$ |
| | | | REACTOME_SRP_DEPENDENT_COTRANSLATIONAL_PROTEIN_TARGETING_TO_MEMBRANE | 85 | 13 | $6.80x10^{-15}$ |
| | | | REACTOME_SELENOAMINO_ACID_METABOLISM | 87 | 13 | $9.32x10^{-15}$ |
| | | | REACTOME_EUKARYOTIC_TRANSLATION_INITIATION | 89 | 13 | $1.27x10^{-14}$ |
| | | | REACTOME_INFLUENZA_INFECTION | 127 | 13 | $1.41x10^{-12}$ |
| | | | REACTOME_RRNA_PROCESSING | 164 | 14 | $2.27x10^{-12}$ |
| | | | REACTOME_REGULATION_OF_EXPRESSION_OF_SLITS_AND_ROBOS | 133 | 13 | $2.56x10^{-12}$ |
| | | | REACTOME_TRANSLATION | 251 | 16 | $4.95x10^{-12}$ |
| | | | REACTOME_SIGNALING_BY_ROBO_RECEPTORS | 177 | 13 | $9.74x10^{-11}$ |

| | | | | | | |
|---|---|---|---|---|---|---|
| | | | REACTOME_DEVELOPMENTAL_BIOLOGY | 833 | 22 | $1.06x10^{-08}$ |
| | | | REACTOME_NERVOUS_SYSTEM_DEVELOPMENT | 522 | 17 | $3.17x10^{-08}$ |
| | | | REACTOME_METABOLISM_OF_AMINO_ACIDS_AND_DERIVATIVES | 316 | 13 | $1.08x10^{-07}$ |
| | | | REACTOME_CELLULAR_RESPONSES_TO_STIMULI | 688 | 18 | $3.22x10^{-07}$ |
| | Oligodendrocytes | Chemical and Genetic pertubation | BLALOCK_ALZHEIMERS_DISEASE_DN | 1171 | 12 | $3.46x10^{-08}$ |
| | | GO cellular component | GOCC_NEURON_PROJECTION | 1241 | 11 | $7.10x10^{-07}$ |
| | | Cell type signature | MURARO_PANCREAS_BETA_CELL | 875 | 11 | $2.02x10^{-08}$ |

**Table S4: Several genes are differentially expressed in medial amygdala cell types in both chronic pain and migraine.** P_bonf = Bonferroni-corrected p value, Z = DEG regression beta value divided by DEG regression standard error.

| | | **Chronic pain** | | | **Migraine** | | |
|---|---|---|---|---|---|---|---|
| Cell | Gene | Z | P | P_bonf | Z | P | P_bonf |
| Endothelial | GLUD1P3 | 6.98 | 3.48E-11 | 6.50E-07 | 4.94 | 1.51E-06 | 2.83E-02 |
| | KDM4B | 5.95 | 1.06E-08 | 1.97E-04 | 4.91 | 1.69E-06 | 3.15E-02 |
| | LIFR | -4.9 | 1.86E-06 | 3.47E-02 | -5.92 | 1.14E-08 | 2.13E-04 |
| | NRBF2 | -4.95 | 1.49E-06 | 2.79E-02 | -6.09 | 4.48E-09 | 8.36E-05 |
| | TNFRSF6B | 5.18 | 4.92E-07 | 9.19E-03 | 4.89 | 1.85E-06 | 3.47E-02 |
| Microglia | CEBPD | 5.21 | 4.14E-07 | 7.73E-03 | 5.19 | 4.52E-07 | 8.45E-03 |
| | MBNL1 | 5.18 | 4.86E-07 | 9.07E-03 | 5.15 | 5.61E-07 | 1.05E-02 |
| | MRPL39 | 4.89 | 1.87E-06 | 3.49E-02 | 5.19 | 4.63E-07 | 8.65E-03 |
| | RBM22 | -5.1 | 6.96E-07 | 1.30E-02 | 4.98 | 1.25E-06 | 2.34E-02 |
| | RPL24 | -5.21 | 4.19E-07 | 7.83E-03 | 4.83 | 2.53E-06 | 4.74E-02 |
| | RPL27 | -5.52 | 9.31E-08 | 1.74E-03 | 4.9 | 1.80E-06 | 3.37E-02 |
| | RRBP1 | 5.04 | 9.28E-07 | 1.73E-02 | 6.31 | 1.40E-09 | 2.61E-05 |

**Table S5: Overlap in migraine and chronic pain DEGs is primarily driven by medial amygdala endothelial cells.** P = hypergeometric test p value.

| Region | Cell type | N DEG overlap | p |
|---|---|---|---|
| dACC | Astrocytes | 0 | 1 |
| | Endothelial | 0 | 1 |
| | ExN | 0 | 1 |
| | InN | 0 | 1 |
| | Microglia | 0 | 1 |
| | Oligodendrocytes | 0 | 1 |
| DLPFC | Astrocytes | 0 | 1 |
| | Endothelial | 0 | 1 |
| | ExN | 0 | 1 |
| | InN | 0 | 1 |
| | Microglia | 0 | 1 |
| | Oligodendrocytes | 0 | 1 |
| BLA | Astrocytes | 0 | 1 |
| | Endothelial | 0 | 1 |
| | ExN | 0 | 1 |
| | InN | 0 | 1 |
| | Microglia | 0 | 1 |
| | Oligodendrocytes | 0 | 1 |
| MeA | Astrocytes | 0 | 1 |
| | Endothelial | 5 | 0.022 |
| | ExN | 0 | 1 |
| | InN | 0 | 1 |
| | Microglia | 7 | 0.16 |
| | Oligodendrocytes | 0 | 1 |

**Table S6: Several genes are differentially expressed in amygdala cell types in both chronic pain and lifetime oxymorphone use.** P_bonf = Bonferroni-corrected p value, Z = DEG regression beta value divided by DEG regression standard error.

| | | | **Chronic pain** | | | **Oxymorphone** | | |
|---|---|---|---|---|---|---|---|---|
| Region | Cell | Gene | Z | P | P_bonf | Z | P | P_bonf |
| BLA | Endothelial | GADD45B | 4.87 | 1.95E-06 | 0.03637 | 6.3 | 1.51E-09 | 2.82E-05 |
| | | YBX3 | 4.81 | 2.63E-06 | 0.049103 | 6.15 | 3.42E-09 | 6.40E-05 |
| | Microglia | CCL2 | 5.7 | 3.49E-08 | 0.000652 | 4.92 | 1.76E-06 | 3.30E-02 |
| | | RASSF10 | -5.36 | 2.02E-07 | 0.003779 | -4.98 | 1.38E-06 | 2.59E-02 |
| | | SERPINA3 | 5.21 | 4.06E-07 | 0.007581 | 4.95 | 1.55E-06 | 2.90E-02 |
| MeA | Endothelial | FAM20C | 5.24 | 3.77E-07 | 0.007041 | 4.95 | 1.44E-06 | 2.68E-02 |
| | | HILPDA | 5.26 | 3.35E-07 | 0.006261 | 4.87 | 2.09E-06 | 3.91E-02 |
| | | KRBA1 | 5.35 | 2.22E-07 | 0.004151 | 5.15 | 5.65E-07 | 1.06E-02 |
| | | LOXL2 | 4.83 | 2.57E-06 | 0.047982 | 4.95 | 1.42E-06 | 2.66E-02 |
| | Microglia | FZD8 | -5.55 | 7.80E-08 | 0.001457 | -4.94 | 1.52E-06 | 2.84E-02 |
| | | RPL27 | -5.52 | 9.31E-08 | 0.001739 | -4.99 | 1.17E-06 | 2.19E-02 |
| | | RPL31 | -6 | 7.44E-09 | 0.000139 | -4.91 | 1.75E-06 | 3.27E-02 |
| | | RPS21 | -5.68 | 3.98E-08 | 0.000744 | -5.49 | 1.05E-07 | 1.97E-03 |

**Table S7: Overlap in lifetime oxymorphone use and chronic pain DEGs is primarily driven by amygdala endothelial cells.** P = hypergeometric test p value.

| Region | Region and cell type | N DEG overlap | p |
|---|---|---|---|
| dACC | Astrocytes | 0 | 1 |
| dACC | Endothelial | 0 | 1 |
| dACC | ExN | 0 | 1 |
| dACC | InN | 0 | 1 |
| dACC | Microglia | 0 | 1 |
| dACC | Oligodendrocytes | 0 | 1 |
| DLPFC | Astrocytes | 0 | 1 |
| DLPFC | Endothelial | 0 | 1 |
| DLPFC | ExN | 0 | 1 |
| DLPFC | InN | 0 | 1 |
| DLPFC | Microglia | 0 | 1 |
| DLPFC | Oligodendrocytes | 0 | 1 |
| BLA | Astrocytes | 0 | 1 |
| BLA | Endothelial | 2 | $9 \times 10^{-04}$ |
| BLA | ExN | 0 | 1 |
| BLA | InN | 0 | 1 |
| BLA | Microglia | 3 | $8 \times 10^{-03}$ |
| BLA | Oligodendrocytes | 0 | 1 |
| MeA | Astrocytes | 0 | 1 |
| MeA | Endothelial | 4 | $2 \times 10^{-04}$ |
| MeA | ExN | 0 | 1 |
| MeA | InN | 0 | 1 |
| MeA | Microglia | 4 | $7 \times 10^{-03}$ |
| MeA | Oligodendrocytes | 0 | 1 |

**Table S8: Genes differentially expressed in lifetime fentanyl use.** pBonferroni = Bonferroni-corrected p value, Z = DEG regression beta value divided by DEG regression standard error. ExN = excitatory neuron, InN = inhibitory neuron.

| **Region** | **cell** | **Gene** | **p** | **$p_{Bonferroni}$** | **Z** |
|---|---|---|---|---|---|
| dACC | Endothelial | PIK3R5 | $1.07x10^{-06}$ | 0.0200 | -5 |
| | ExN | PIK3R5 | $4.21x10^{-07}$ | $7.88x10^{-03}$ | -5.195 |
| | InN | PIK3R5 | $1.97x10^{-06}$ | 0.0367 | -4.876 |
| BLA | Endothelial | EXTL3 | $1.53x10^{-06}$ | 0.0286 | 4.935 |
| | | PHLPP2 | $1.87x10^{-06}$ | 0.0350 | 4.892 |
| | | GSK3B | $2.41x10^{-06}$ | 0.0451 | 4.836 |
| | | NUP50 | $9.66x10^{-07}$ | 0.0181 | 5.033 |
| | | MS4A4A | $8.41x10^{-07}$ | 0.0157 | -5.063 |
| | | ASTN1 | $1.66x10^{-06}$ | 0.0311 | 4.917 |
| | | GNE | $4.83x10^{-07}$ | $9.03 x10^{-03}$ | 5.179 |
| | | HPGDS | $8.35x10^{-07}$ | 0.0156 | -5.064 |
| | | TCEAL7 | $2.32x10^{-07}$ | $4.32 x10^{-03}$ | -5.331 |
| | | CDRT15 | $6.43x10^{-08}$ | $1.20 x10^{-03}$ | -5.588 |
| | ExN | ZNF806 | $5.03x10^{-07}$ | $9.39 x10^{-03}$ | -5.164 |
| | | YWHAQ | $8.70x10^{-07}$ | 0.0163 | -5.049 |
| | | AKAP17A | $1.68x10^{-07}$ | $3.13 x10^{-03}$ | -5.389 |
| MeA | Astrocytes | LINC00863 | $5.16x10^{-07}$ | $9.64 x10^{-03}$ | -5.213 |
