## Supplementary material for "The impact of chronic pain on brain gene expression": Table1

**Demographic information, VA National PTSD Brain Bank donors**. PMI = Post Mortem Interval. Oxymorphone = lifetime oxymorphone use, fentanyl = lifetime fentanyl use.

| **Trait** | **Trait Status** | **Mean age at death** | **Mean PMI** | **Female N (%)** | **Total N** |
| --- | --- | --- | --- | --- | --- |
| Chronic Pain | 0 | 45.08 | 29.05 | 74 (34%) | 220 |
|  | 1 | 46.36 | 27.21 | 41 (49%) | 84 |
| Migraine | 0 | 46.34 | 28.33 | 92 (36%) | 256 |
|  | 1 | 40.63 | 29.67 | 23 (48%) | 48 |
| Oxymorphone | 0 | 45.47 | 28.66 | 106 (37%) | 287 |
|  | 1 | 44.94 | 26.65 | 9 (53%) | 17 |
| Fentanyl | 0 | 46.02 | 28.87 | 99 (37%) | 270 |
|  | 1 | 40.75 | 26.11 | 15 (47%) | 32 |
|  | NA | 41.01 | 23.75 | 1 (50%) | 2 |
