## Supplementary material for "The impact of chronic pain on brain gene expression": Table2

**Testing for enrichment of chronic pain genes in mouse brain and nerve tissue**. Fisher’s test p values from tests of enrichment of chronic pain genes (per tissue and cell type) within mouse brain and nerve tissue transcriptomics results for pain experiments (McGill TSPdb).

| **Region** | **Cell type** | **Spinal ganglia** | **microglia** | **Sciatic nerve** | **Brain stem** |
| --- | --- | --- | --- | --- | --- |
| dACC | Astrocytes | 1 | 1 | 0.17 | 1 |
|  | Endothelial | 1 | 0.00 | 1 | 1 |
|  | ExN | 0.64 | 1 | 0.23 | 1 |
|  | InN | 0.56 | 1 | 0.14 | 1 |
|  | Microglia | 0.46 | 0.01 | 0.16 | 1 |
|  | Oligodendrocytes | 0.35 | 1 | 1 | 1 |
| DLPFC | Astrocytes | 1 | 1 | 1 | 1 |
|  | Endothelial | 0.33 | 1 | 1 | 1 |
|  | ExN | 1 | 1 | 1 | 1 |
|  | InN | 1 | 1 | 1 | 1 |
|  | Microglia | 1 | 1 | 1 | 1 |
|  | Oligodendrocytes | 1 | 1 | 1 | 1 |
| BLA | Astrocytes | 1 | 1 | 1 | 1 |
|  | Endothelial | 1 | 1 | 1 | 1 |
|  | ExN | 1 | 1 | 1 | 1 |
|  | InN | 1 | 1 | 1 | 1 |
|  | Microglia | 0.16 | 0.05 | 0.11 | 1 |
|  | Oligodendrocytes | 0.33 | 1 | 1 | 1 |
| MeA | Astrocytes | 0.26 | 0.01 | 0.23 | 1 |
|  | Endothelial | 0.93 | 0.05 | 0.70 | 1 |
|  | ExN | 0.46 | 1 | 1 | 1 |
|  | InN | 1 | 1 | 1 | 1 |
|  | Microglia | 0.01 | 1 | 0.77 | 1 |
|  | Oligodendrocytes | 0.65 | 1 | 0.23 | 1 |
