## Supplementary material for "The impact of chronic pain on brain gene expression": Table3

**Comparing microglia DEGs to DAM/ARM Genes.** Chronic pain microglia DEGs found to be up or down-regulated in DAMs and/or ARMs. Z = our DEG analysis beta / SE.

| **Region** | **Gene name** | **Z** | **KerenShaul_DAM** | **SalaFrigerio_ARM** |
| --- | --- | --- | --- | --- |
| BLA | ADAP2 | -5.83 | . | Down |
|  | ADSSL1 | 6.498 | Up | Up |
|  | ANKRD55 | -5.285 | Up | . |
|  | ATF3 | 5.277 | Up | Up |
|  | ATOX1 | -4.949 | Up | . |
|  | BCL2A1 | 4.966 | . | Up |
|  | CD164 | 4.888 | Down | Down |
|  | FAU | -6.61 | Up | Up |
|  | FLT1 | 6.718 | Up | . |
|  | GCNT2 | -6.231 | . | Up |
|  | GRN | -4.972 | Up | . |
|  | HIF1A | 6.211 | Up | Up |
|  | LDHA | 5.411 | Up | Up |
|  | MAFF | 6.155 | Up | . |
|  | MGAT4A | 5.232 | . | Down |
|  | NRP1 | 5.467 | Up | . |
|  | OLFML3 | -6.164 | . | Down |
|  | PFDN5 | -5.471 | Up | . |
|  | PGK1 | 5.726 | Up | . |
|  | PLAUR | 5.44 | Up | Up |
|  | RAMP1 | -5.089 | Up | Up |
|  | RHOB | 4.97 | Down | Down |
|  | RPL13A | -5.665 | Up | . |
|  | RPL18 | -5.831 | Up | . |
|  | RPL19 | -5.568 | . | Up |
|  | RPL3 | -5.315 | Up | . |
|  | RPL32 | -6.286 | Up | Up |
|  | RPL4 | -5.262 | Up | . |
|  | RPL41 | -5.408 | . | Up |
|  | RPL6 | -5.179 | Up | . |
|  | RPL7 | -5.009 | Up | . |
|  | RPL8 | -5.609 | Up | . |
|  | RPLP1 | -6.13 | Up | Up |
|  | RPS11 | -5.146 | Up | . |
|  | RPS14 | -5.4 | Up | Up |
|  | RPS16 | -5.54 | Up | Up |
|  | RPS18 | -5.123 | Up | Up |
|  | RPS3 | -6.556 | Up | . |
|  | RPS4X | -6.209 | Up | Up |
|  | RPS5 | -5.935 | Up | Up |
|  | RPS9 | -5.137 | Up | . |
|  | SALL1 | -4.845 | . | Down |
|  | SCAMP2 | -5.771 | . | Down |
|  | SLC11A1 | 6.67 | Up | Up |
|  | SLC16A3 | 6.479 | . | Up |
|  | SLC2A1 | 6.13 | Up | Up |
|  | SLC2A5 | 5.084 | . | Down |
|  | SPP1 | 5.559 | Up | Up |
|  | SSR4 | -6.116 | Up | . |
|  | SUSD3 | -5.587 | . | Down |
|  | TGFBR1 | 6.185 | . | Down |
|  | TLR2 | 6.27 | Up | Up |
|  | TMEM119 | -5.638 | Down | Down |
|  | TREM2 | -5.122 | Up | Up |
|  | USE1 | -6.724 | Up | . |
| dACC | CD84 | -5.296 | Up | . |
|  | SELPLG | -5.854 | Down | Down |
|  | SUSD3 | -5.851 | . | Down |
| MeA | ATP6V0E1 | -5.104 | Up | . |
|  | RPL32 | -5.061 | Up | Up |
|  | RPL7 | -4.997 | Up | . |
|  | RPS12 | -4.955 | Up | Up |
|  | RPS21 | -5.682 | Up | Up |
|  | RPS3 | -5.33 | Up | . |
|  | SLC11A1 | 5.585 | Up | Up |
|  | TGFBR1 | 5.047 | . | Down |
|  | TLR2 | 5.253 | Up | Up |
