## Supplementary material for "The impact of chronic pain on brain gene expression": Table4

**Genes associated with predisposition to chronic pain (**PRS-DEGs). P_bonf = Bonferroni-adjusted P value

| **Region** | **Cell type** | **Gene Name** | **p** | **p_bonferroni_** | **Z** |
| --- | --- | --- | --- | --- | --- |
| dACC | Oligodendrocytes | FAM13B | 2.34x10^-06^ | 0.0436 | -4.879 |
|  |  | FAM184B | 1.12X10^-07^ | 0.0021 | 5.528 |
|  |  | CDH17 | 2.67X10^-06^ | 0.0499 | 4.849 |
|  |  | TRAF5 | 4.48X10^-08^ | 0.0008 | 5.715 |
|  |  | DOCK3 | 2.54X10^-06^ | 0.0475 | 4.86 |
|  |  | LAMB1 | 8.77X10^-08^ | 0.0016 | 5.579 |
|  |  | PLXDC2 | 5.86X10^-07^ | 0.0109 | -5.182 |
|  |  | DUSP4 | 1.65X10^-06^ | 0.0308 | -4.956 |
|  |  | NCOA5 | 6.59X10^-09^ | 0.0001 | -6.092 |
|  |  | EPHA5 | 1.96X10^-07^ | 0.0037 | 5.413 |
|  |  | SCUBE3 | 2.57X10^-06^ | 0.0480 | 4.858 |
|  |  | PFKFB1 | 2.45X10^-07^ | 0.0046 | 5.367 |
|  |  | ANO10 | 1.06X10^-06^ | 0.0198 | 5.054 |
|  |  | C1QTNF7 | 2.07X10^-08^ | 0.0004 | 5.869 |
|  |  | NFKBID | 2.51X10^-07^ | 0.0047 | -5.361 |
|  |  | FZD4 | 6.33X10^-07^ | 0.0118 | 5.165 |
|  |  | MRPL48 | 1.15X10^-06^ | 0.0214 | -5.037 |
|  |  | PDXDC2P | 1.80X10^-06^ | 0.0336 | 4.938 |
|  |  | RP11.362A1.1 | 2.62X10^-06^ | 0.0489 | 4.854 |
| MeA | Microglia | ACCS | 1.85X10^-06^ | 0.0346 | -4.92 |
|  |  | GPR173 | 2.47X10^-06^ | 0.0462 | 4.856 |
